# NEC-Associated DNA Methylation Signatures in Colon are Evident in Stool Samples of Affected Individuals

**DOI:** 10.1101/2020.11.30.20241307

**Authors:** Misty Good, Tianjiao Chu, Patricia Shaw, Lila S. Nolan, Lora McClain, Austin Chamberlain, Carlos Castro, Qingqing Gong, Krista Cooksey, Laura Linneman, David N. Finegold, David Peters

## Abstract

Neonatal necrotizing enterocolitis (NEC) is a devastating and unpredictable gastrointestinal disease with a high mortality rate in premature infants. Currently, no predictive or diagnostic biomarkers exist for NEC. Clinical intervention is reactive to the overt manifestations of disease resulting in high levels of morbidity and mortality. To better understand the molecular mechanisms that underpin NEC, we have undertaken a high resolution genome wide epigenomic analysis using solution phase hybridization and next generation DNA sequencing of bisulfite converted DNA. Our data reveal a broad and significant genomic hypermethylation in surgical NEC tissues compared to non-NEC controls. These changes were found to be far more pronounced in regions outside CpG islands and gene regulatory elements, which suggests that NEC-specific hypermethylation is not a non-specific global phenomenon. We identified a number of important biological pathways that are dysregulated in NEC and observed a clear association between NEC methylation changes and gene expression. Significantly, we found that the same patterns of global methylation identified in surgical NEC tissue are also detectable in stool samples from affected infants. To our knowledge, this is the first evidence of a methylomic signature that is both associated with NEC and detectable non-invasively. These findings point towards a new opportunity for the development of novel screening, diagnostic and phenotyping methods for NEC that could be deployed in the NICU for improved detection of this devastating disease.

## Introduction

Necrotizing enterocolitis (NEC) is a deadly gastrointestinal disease that primarily affects premature infants for which there is no clinical biomarker available. This is a major challenge in the neonatal intensive care unit (NICU) not having the ability to determine which babies are at the highest risk for NEC, obstructing a path to identify a window for preventative strategies. NEC affects ∼8% of all premature infants and is the most common cause of death in infants from 15 to 60 days after birth^1^. The presentation of NEC can either be acute or insidious with the presence of abdominal distension, feeding intolerance and bloody stools that may quickly progress to the need for surgical removal of necrotic intestine^2^. The search for potential non-invasive biomarkers is important as careful monitoring of these markers in the stool could predict a NEC diagnosis and afford the ability to intervene earlier in the hospital course.

The ability to identify disease-specific cell-free nucleic acid signatures in plasma, tissues other fluid reservoirs including stool has been demonstrated^3-5^. Cell-free DNA as a non-invasive biomarker can assess an ongoing inflammatory state and detect cellular death^6,7^. In this regard, the clinical utility of genomic sequencing of DNA during routine clinical care is now well established in specific patient populations^8,9^. These transformative developments in non-invasive plasma DNA analysis revolutionized the standard of prenatal care. Epigenetic modification, such as DNA methylation, has several features that make it an attractive target for the development of biomarkers for diagnosis and phenotyping in precision medicine. For example, DNA methylation signatures convey rich and detailed insight into the molecular phenotype of their cell type/tissue of origin^10^. One such example is DNA methylation profiling in inflammatory bowel disease, where several investigators have evaluated intestinal, whole blood and B cell specific methylomes during active ulcerative colitis or Crohn’s disease flares^11-15^. Alterations in the epigenomic molecular phenotype at the level of DNA methylation are intimately associated with pathogenesis of disease and exposure to environmental stressors. However, in the context of NEC, the methylation patterns in stool samples of premature infants have not been evaluated.

In this study, we sought to determine whether these innovations in epigenomics are applicable to the neonatal setting to enable the development of screening methods for NEC and to improve our understanding of NEC pathogenesis. We now hypothesize that the stool of infants will exhibit a distinct signature at the onset of NEC and that the colonic tissue methylome shares features with the stool methylation signature. To test our hypotheses, we performed a high resolution genome-wide epigenomic analysis of NEC tissue using solution phase hybridization and next generation DNA sequencing of bisulfite converted DNA. Similar analysis was performed on stool samples obtained from infants with NEC and from healthy controls. Our findings reveal that colonic NEC samples are broadly, yet selectively, hypermethylated in NEC tissue compared with non-NEC controls. We found that NEC is associated with alterations in the methylation patterns of key DNA methylases. We also describe the relationship between DNA methylation and RNA transcription in colon resected for NEC. Importantly, we discovered that NEC-associated hypermethylation is also evident in stool samples from infants with NEC. Our results demonstrate that selective genome-wide DNA hypermethylation in the both tissue and stool is a striking feature of individuals affected by NEC. These findings suggest that epigenomic liquid biopsy of stool may provide a novel screening and/or phenotyping approach for this devastating disease.

## Methods

### Study Population and Selection Criteria

#### Intestine

The intestinal samples in this study were collected in accordance with the University of Pittsburgh anatomical tissue procurement guidelines and was approved by the University of Pittsburgh Institutional Review Board (IRB) Protocols (PRO09110437 or PRO14070508). Premature infants were recruited under Protocol PRO09110437 at either Children’s Hospital of Pittsburgh (CHP) of University of Pittsburgh Medical Center (UPMC) or Magee-Womens Hospital Neonatal Intensive Care Units (NICUs) and consent was obtained their parent or legal guardian. Intestinal tissue samples were procured if resected for NEC or other non-inflammatory indications, such as reanastomosis, spontaneous intestinal perforation or anorectal malformation. In some instances, deidentified intestinal samples were obtained with a waiver of consent and approval of University of Pittsburgh IRB (PRO14070508). For these cases, the clinical information obtained was limited to the location of the resected intestinal tissue sample and the surgical indication. Resections of intestinal tissue were snap frozen and stored at −80 degrees Celsius until further analysis. Non-NEC tissue samples are healed NEC tissue obtained during surgical re-anastomosis. These samples are a readily available source of human tissue for this study design and the practice of using these as controls is standard in this field. Due to the manner in which samples were obtained prior to the onset of this study, we were unable to obtain complete information regarding neonatal sex for all surgically resected samples. In this regard, we have therefore focused only on autosomal data in the evaluation of the intestine to avoid the complication of X-inactivation and minimize the impact of sex differences.

#### Stool

Stool samples were obtained after informed consent was obtained from the parents on behalf of their infants admitted to St. Louis Children’s Hospital Neonatal Intensive Care Unit after IRB approval from Washington University in St. Louis (Protocol Number 201706182). Stool was collected from diapers and identified as cases with NEC or premature control infants (as in Table S9). The diapers were initially placed at 4 degrees Celsius, transferred to the laboratory within 12-24 hours, aliquoted into barcoded tubes and stored at −80 degrees Celsius until processing.

### DNA Recovery from Tissue Sections

Snap frozen specimens were mounted on appropriate embedding molds (Large, Thermo Scientific #2219; or Small, Sakura Tissue - Tek #4566) with clear OCT compound (Optimal Cutting Temperature Embedding Medium) (Fisher HealthCare #4585), and sectioned with a cryostat (Leica CM 1850 UV), 7 microns). These sections were mounted on membrane slides (Leica PEN - Membrane Slide, 2.0 microns #11505158), stained with toluidine blue (Toluidine Blue 0.1% Aqueous, Newcomer Supply #14027), and air dried (Sampla Dry Keeper, Samplatec. Corp). DNA was extracted from tissue sections using the Nucleospin Tissue XS Kit (Macherey-Nagel). Extracted DNA was quantified by using the KAPA hgDNA Quantification and QC Kit (Roche).

### DNA Recovery from Stool

Stool DNA was extracted using NucleoSpin DNA Stool, Mini kit (Macherey-Nagel). Human and bacterial DNA content were measured using Femto Human and Femto Bacterial DNA Quantification Kits (Zymo).

### Bisulfite Sequencing

Stool and tissue DNA were sheared with Covaris to a size of ∼175bp. Libraries were prepared using the KAPA HyperPrep Kit (Roche). Libraries were bisulfite converted post-ligation using the EZ DNA Methylation-Direct Kit (Zymo) and amplified 13-15 cycles. Bisulfite-converted DNA libraries underwent targeted capture using the SeqCap Epi CpGiant Enrichment Kit (Roche, Pleasanton, CA) and amplified 10 cycles. Samples were sequenced on a HiSeq 2500 (Illumina) with 100bp paired-end reads to a mean targeted read depth of ∼24x. DNA sequence reads were quality trimmed and adaptor sequences were removed using Trim-Galore (https://www.bioinformatics.babraham.ac.uk/projects/trim_galore/). The reads were aligned to the human reference sequence (GRCh38/hg20) using Bisulfite Read Mapper^16^ in paired-end, bowtie2 mode. Any unmapped reads were aligned in single-end, bowtie2 mode. Read duplicates were removed using Bismark. Methylation was called on paired-end and single-end files and then merged to determine the methylation status for each CpG site. DNA methylation signatures were identified using the beta-binomial test implemented in the R packages methylSig^17^ and DSS^18^.

## Results

Targeted genome-wide bisulfite sequencing was carried out on DNA extracted from histological tissue sections obtained from NEC colon (n=8) and non-NEC colon (n=13). A total of 2,069,470,947 aligned read pairs across all samples were sequenced. Solution-phase targeted bisulfite DNA sequencing was used, as this method delivers high sequencing read depth at single base resolution in well-characterized regions of the human genome including promoters, exons, introns, CpG islands, CpG island shores and enhancers. The capture regions include ∼5.5 million CpG sites. This approach quantifies DNA methylation levels within genes involved in known biological pathways.

### Colonic NEC Samples are Broadly but Selectively Hypermethylated

We first explored broad changes in DNA methylation between NEC and non-NEC colon samples. Multidimensional scaling revealed a general separation between colon NEC and non-NEC samples (Figure 1A). As shown in Figure 1B and Table 1, we found that CpGs in NEC samples were frequently more methylated than their non-NEC counterparts. Notably, these effects were largely confined to regions without CGIs, whereas in CGIs outside promoters, we found that DNA methylation was generally more balanced between diseased and control samples. As an exception to this broadly hypermethylated state, CGIs within promoters of non-NEC control samples were generally more methylated than their NEC counterparts. Thus, the genomes of NEC colon samples exist in a broadly hypermethylated state relative to their non-NEC counterparts. However, this effect was largely observed outside of CGIs and, in contrast, CGIs in promoter regions were hypomethylated in NEC compared to non-NEC colon.

**Table 1:** Proportion of CpG sites in different genomic elements that were hyper- or hypomethylated in NEC versus non-NEC control colon.

|  | Non-NEC | NEC | Chi-square p-value<br>versus "All Sites"<br>Distribution |
| --- | --- | --- | --- |
| All Sites | 1046175 (41.82%) | 1455607 (58.18%) |  |
| Promoter No CGI | 149134 (41.98%) | 206095 (58.02%) | p-value = 0.06188 |
| Promoter & CGI | 244415 (56.40%) | 188912 (43.60%) | p-value < 0.05 |
| Intron/Exon No CGI | 276685 (35.06%) | 512492 (64.94%) | p-value < 0.05 |
| Intergenic CGI | 61966 (51.60%) | 58125 (48.40%) | p-value < 0.05 |
| Intron/Exon & CGI | 275356 (50.12%) | 274061 (49.88%) | p-value < 0.05 |
| CGI Shores | 43281 (45.79%) | 51244 (54.21%) | p-value < 0.05 |
| Intergenic | 154230 (34.87%) | 288112 (65.13%) | p-value < 0.05 |

**Figure 1:**
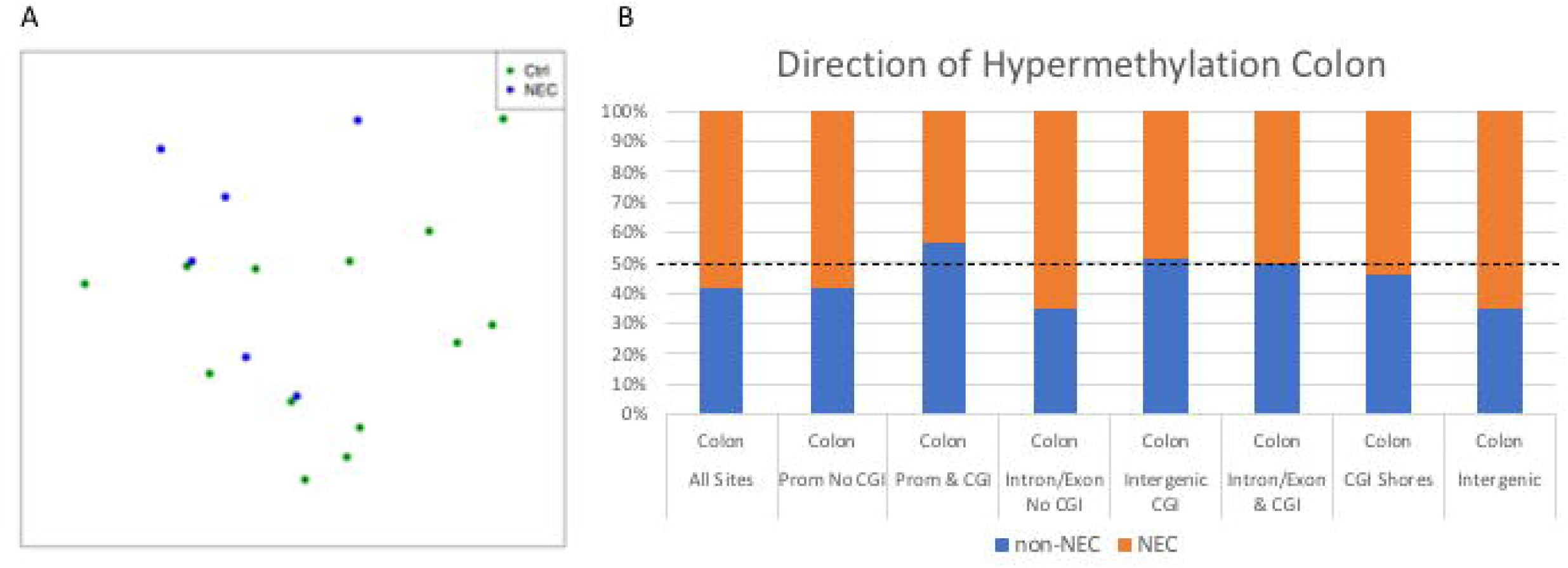
(A) Multi-dimensional scaling (Eucladian) analysis of NEC (blue) and non-NEC (green) colon samples. (B) Proportion of CpG sites in different genomic elements that were hypermethylated in NEC (orange) versus hypermethylated in non-NEC control (blue) in colon. The dotted line represents a 50% ratio of hypermethylated sites in either type of tissues in a given genomic element.

Density plot analysis of all targeted sites revealed a bimodal distribution of CpG methylation with one peak near 0% methylation and another peak approaching 100% methylation (Figure 2A). The relative distribution varied dramatically between different genomic regions. For example, introns/exons and intergenic regions not containing CGIs displayed a considerable shift towards a hypermethylated state (Figure 2D and H) whereas promoter regions not overlapping with CGIs and CGIs outside promoter regions contained considerably more hypomethylated CpG sites. CGIs within promoters were found to be almost completely hypomethylated. Though these patterns of CpG methylation were very similar between both NEC and non-NEC samples, clear differences were apparent. NEC-specific increases in CpG methylation were observed in varying degrees in all genomic contexts (Figure 2 and Table 2). Further examination revealed that CpG sites could be grouped into low methylation (LM) (<20%), intermediate methylation (IM) (20-80%) or high methylation (HM) (>80%) categories (Figure 3). These distributions were altered between NEC and non-NEC colon samples to a degree that reached statistical significance in every case (Table 3). Specifically, hypermethylation in NEC colons was clearly evident and appeared to result from a relative shift from 20-80% CpG methylation to >80% methylation (Figure 3 and Table 3).

**Table 2:** Differences in distributions of DNA methylation (plotted in Figure 2A) between NEC and non-NEC samples within defined genomic regions classes (*a:* median difference, *b:* mean difference, *c:* adjusted for multiple comparisons using Holm’s method)

| Region | diff.med <sup>a</sup> | diff.mean <sup>b</sup> | w.holm <sup>c</sup> |
| --- | --- | --- | --- |
| A: All sites | 0.12478411 | 0.72317302 | p=<0.05 |
| B: Promoter no CGI | 0.11941805 | 0.81540044 | p=<0.05 |
| C: Promoter with CGI | -0.0243087 | 0.0324467 | p=<0.05 |
| D: Intron/exon no CGI | 0.76086895 | 1.06709367 | p=<0.05 |
| E: Intergenic with CGI | -0.0192987 | 0.18622534 | p=<0.05 |
| D: Intron/exon with CGI | -0.0009022 | 0.24585563 | p=<0.05 |
| G: CGI Shores | 0.16172223 | 0.19984825 | p=<0.05 |
| H: Intergenic | 0.72489659 | 1.16574872 | p=<0.05 |

**Table 3:** Distributions of low methylation (LM), intermediate methylation (IM) and high methylation (HM) in genetic elements (NEC vs. non-NEC).

|  |  | LM<br>≤20% | IM >20<br>and<br><80% | HM<br>≥80% | Chi-square p-value<br>non-NEC v. NEC |
| --- | --- | --- | --- | --- | --- |
| All Sites | Non-NEC Colon | 920116<br>(36.20%) | 650160<br>(25.58%) | 971756<br>(38.23%) | p-value < 2.2e-16 |
|  | NEC Colon | 925350<br>(36.40%) | 590865<br>(23.24%) | 1025817<br>(40.35%) |  |
| Promoter<br>No CGI | Non-NEC Colon | 172071<br>(47.72%) | 114283<br>(31.69%) | 74259<br>(20.59%) | p-value < 2.2e-16 |
|  | NEC Colon | 171637<br>(47.60%) | 106225<br>(29.46%) | 82751<br>(22.95%) |  |
| Promoter &<br>CGI | Non-NEC Colon | 428306<br>(93.93%) | 15450<br>(3.39%) | 12227<br>(2.68%) | p-value = 6.028e-15 |
|  | NEC Colon | 428622<br>(94.00%) | 14347<br>(3.15%) | 13014<br>(2.85%) |  |
| Intron/Exon<br>No CGI | Non-NEC Colon | 79563<br>(10.04%) | 267556<br>(33.75%) | 445694<br>(56.22%) | p-value < 2.2e-16 |
|  | NEC Colon | 80883<br>(10.20%) | 241752<br>(30.49%) | 470178<br>(59.31%) |  |
| Intergenic<br>CGI | Non-NEC Colon | 68869<br>(56.41%) | 21603<br>(17.69%) | 31625<br>(25.90%) | p-value < 2.2e-16 |
|  | NEC Colon | 69324<br>(56.78%) | 19843<br>(16.25%) | 32930<br>(26.97%) |  |
| Intron/Exon<br>& CGI | Non-NEC Colon | 389101<br>(68.47%) | 51460<br>(9.06%) | 127719<br>(22.47%) | p-value < 2.2e-16 |
|  | NEC Colon | 391126<br>(68.83%) | 44729<br>(7.87%) | 132425<br>(23.30%) |  |
| CGI<br>Shores | Non-NEC Colon | 28579<br>(30.05%) | 32757<br>(34.44%) | 33768<br>(35.51%) | p-value < 2.2e-16 |
|  | NEC Colon | 29757<br>(31.29%) | 29620<br>(31.14%) | 35727<br>(37.57%) |  |
| Intergenic | Non-NEC Colon | 32655<br>(7.35%) | 156846<br>(35.29%) | 254922<br>(57.36%) | p-value < 2.2e-16 |
|  | NEC Colon | 33043<br>(7.44%) | 143542<br>(32.30%) | 267838<br>(60.27%) |  |

**Figure 2:**
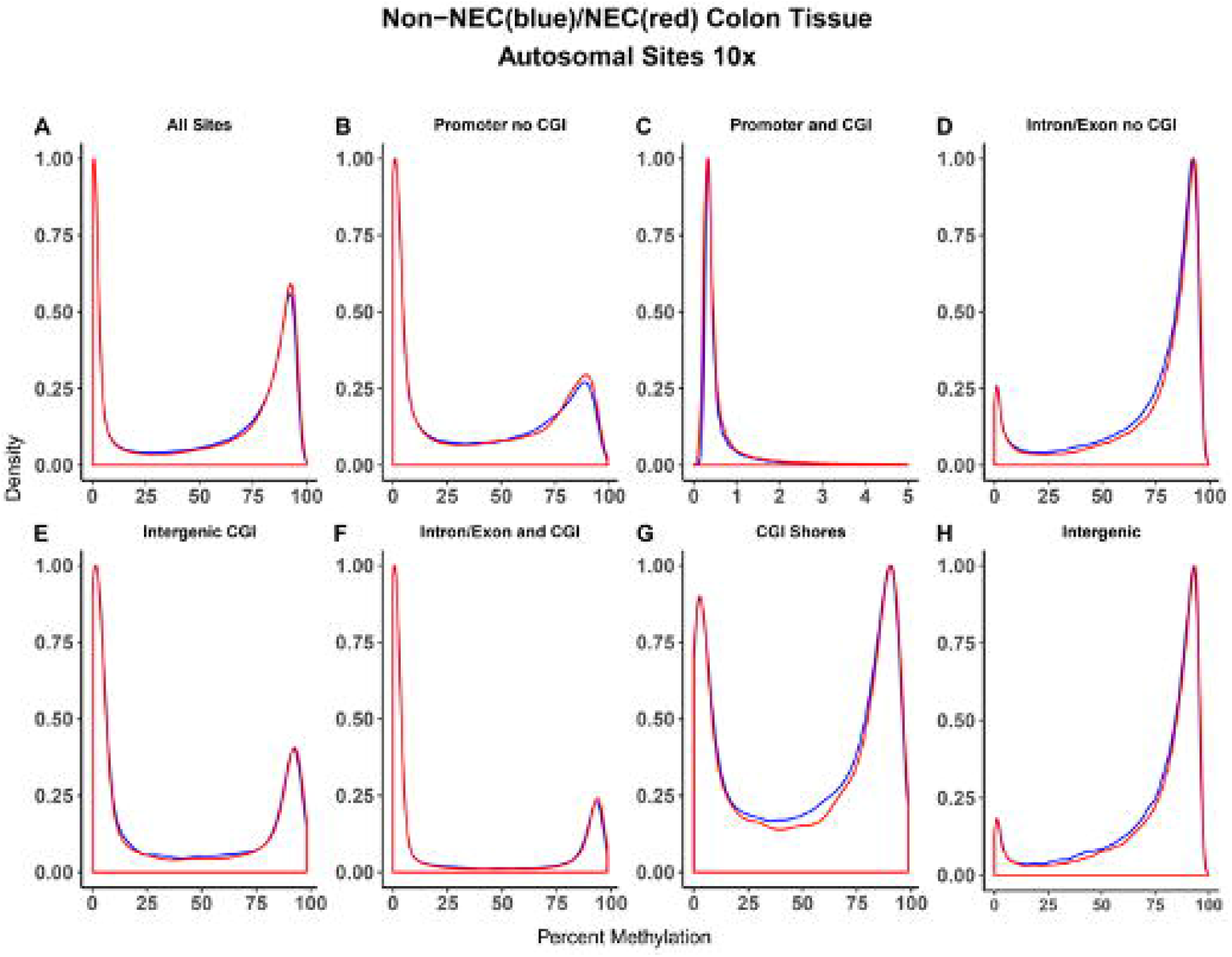
Distribution of methylation of non-NEC (blue) versus NEC (red) colon by genomic element.

**Figure 3:**
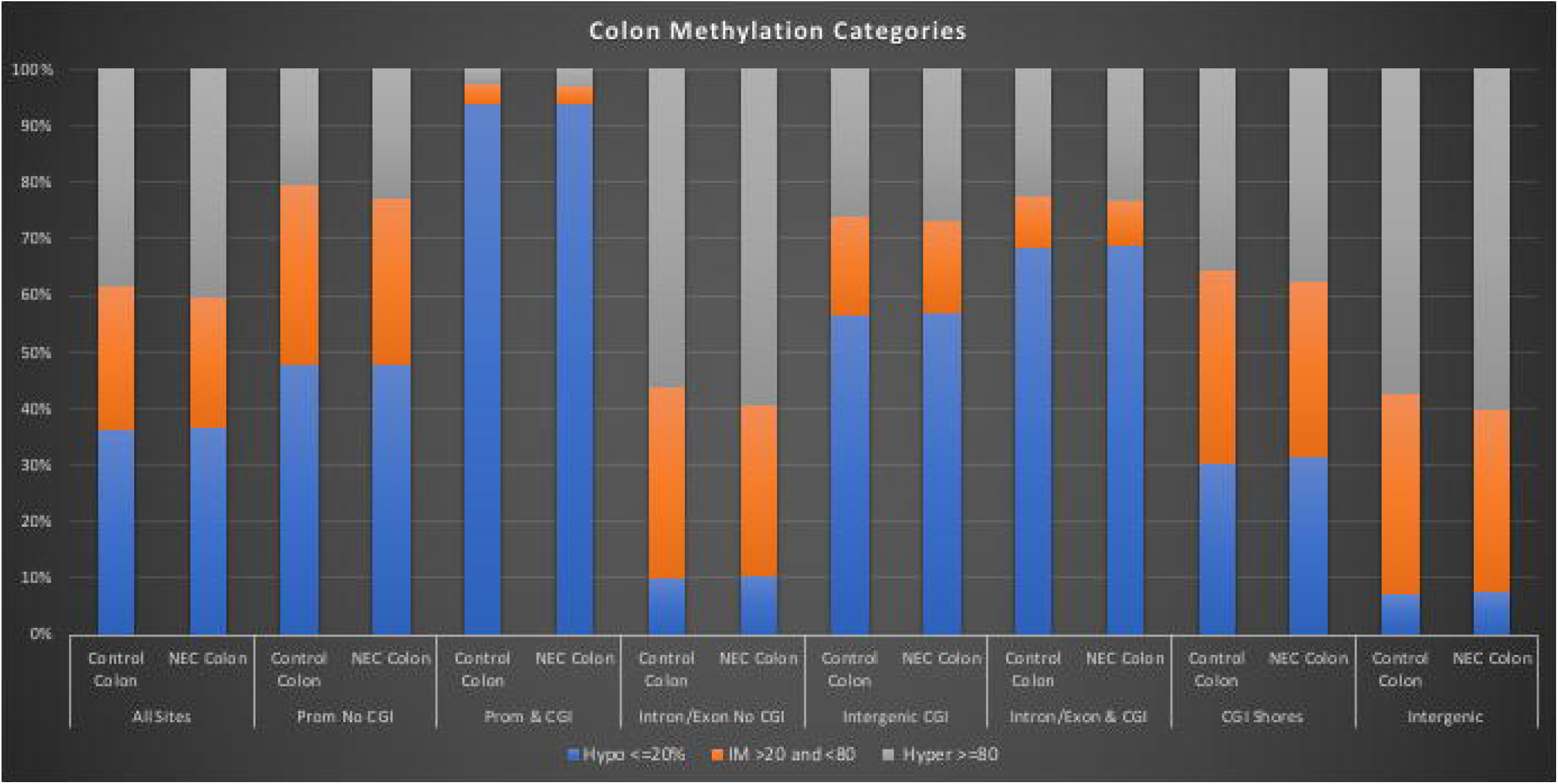
Comparison methylation category distribution (Hypo ≤20% Blue, IM >20% and <80% Orange, Hyper ≥80% Gray) of non-NEC and NEC colon by genomic element.

We also spatially mapped CpG methylation with the goal of visualizing DNA methylation differences between NEC and non-NEC samples across each autosome. These analyses confirmed the relative global hypermethylation of NEC colon when compared to non-NEC controls in regions that do not overlap with CGIs. This approach also confirmed that, at this broad resolution, CpG methylation within CGIs was least affected by NEC regardless of CGI location (Figure 4).

**Figure 4:**
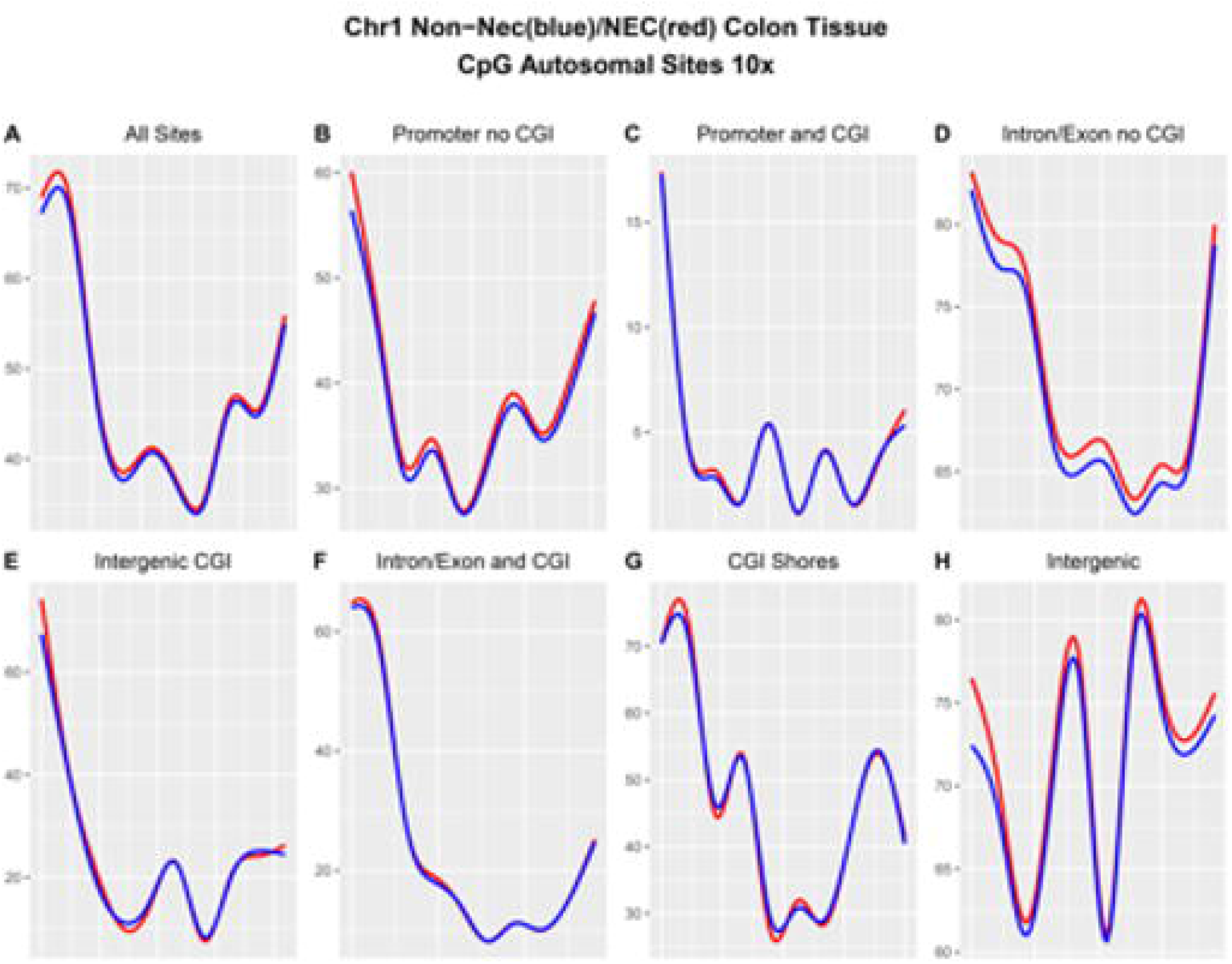
Non-NEC (blue) and NEC (red) methylation across chromosome 1 broken down by genomic element.

### NEC is Associated with Alterations in the Methylation Patterns of Key DNA Methylases

As NEC colon tissues are broadly hypermethylated relative to their non-NEC counterparts, we then explored the methylation levels of genomic regions surrounding genes that are known to influence the epigenome at the level of DNA methylation. We found notable changes in NEC colon tissue relative to the controls. Specifically, we observed a broad reduction in CpG methylation in NEC colon DNA within introns 4-6 (region 2) of DNA Methyltransferase 3a (DNMT3A) (Figure S1A). This region contains a CGI and its adjacent shores and overlaps with an enhancer element(s) and the coding sequence of microRNA 1301 (MIR1301) (Table S1A). We also identified increased NEC-specific CpG methylation within a promoter/enhancer CGI (region 1) of DNMT3A (Figure S1, Table S1A). In DNMT3B, we identified a NEC-specific reduction in DNA methylation within the promoter region (Figure S1B, Table S1B). We also found a NEC-specific reduction in CpG methylation within a promoter/enhancer (region 1) of Tet Methylcytosine Dioxygenase 1 (TET1) and NEC-specific increases in CpG methylation within exon 2 (region 2) and intron 3 (region 3) (Figure S1C, Table S1D). TET3 was found to be hypomethylated in NEC colon samples within a proximal promoter/enhancer region and also a distal enhancer. Regions of NEC hypermethylation were found within introns 1-3 of TET3 (Figure S1D, Table S1D). Notably, we also found that the DNA methylation machinery component, ubiquitin-like protein containing PHD and RING finger domains 1 (UHRF1) was hypomethylated within its promoter region in NEC colon (Figure S1E, Table S1E). These observations suggest a fundamental alteration in the DNA methylation levels of genes encoding proteins that affect dynamic methylation states in differentiated cells.

### Confirmation of DNA Methylation Changes

We have previously undertaken whole genome bisulfite sequencing (WGBS) analysis of enterocytes obtained via laser capture microdissection from surgically resected NEC and non-NEC colon ^19^. These data were compared to our current analysis to estimate the degree of correlation. As shown in Figure 5, when comparing site-specific CpG methylation levels of all targeted sites between NEC colon samples analyzed by WGBS after laser capture with the data in the current study, we found a correlation coefficient of 0.977. For non-NEC controls, the value was 0.962 (Figure 5A). When we only considered sites with a methylation difference of >5% (in either way) between NEC and non-NEC and test p value <0.05 in both data set (laser capture/WGBS and current data) the correlation coefficients were 0.926 between NEC samples and 0.742 between non-NEC samples (Figure 5B). We next compared the degree of CpG site-specific differential methylation between the two data sets and found a correlation coefficient of 0.75 when all sites were considered (Figure 5C) and 0.94 when only sites with a methylation difference >5% (in either way) and test p value <0.05 were considered (Figure 5D). These results demonstrate that there is a considerable degree of correlation between the targeted genome-wide data presented herein and those data obtained following laser capture microdissection of enterocytes and WGBS.

**Figure 5:**
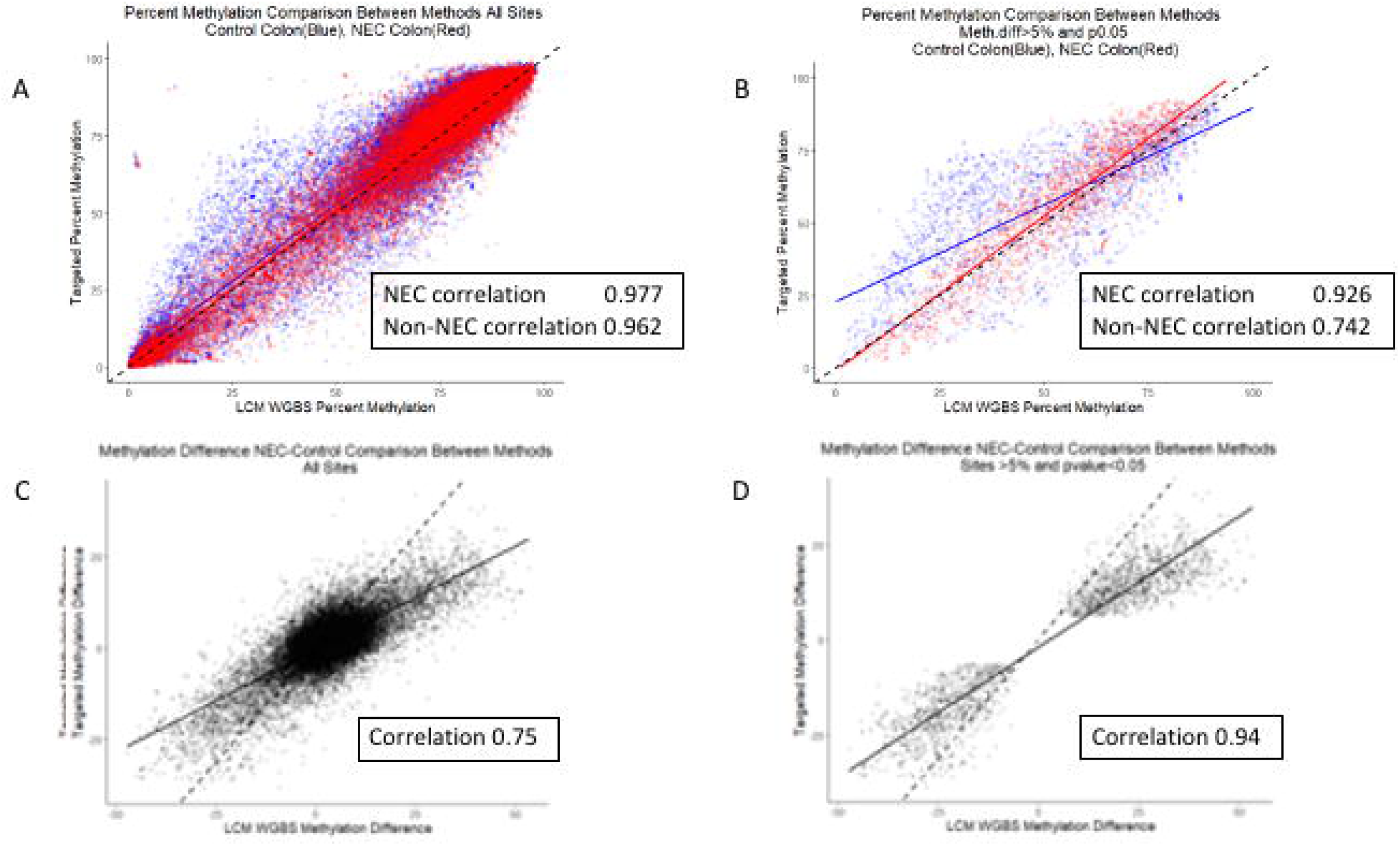
(A and B) Comparison of percent methylation of NEC (red) and non-NEC (blue) between current data from colon tissue and previously published WGBS data of Laser Capture Microscopy of enterocytes^19^. (A) All sites shared. (B) Shared sites with a minimum absolute methylation difference of 5% or more between NEC and non-NEC and p<0.05 in both methods. (C and D) Comparison of methylation difference (NEC minus non-NEC) between colon tissue and WGBS of LCM enterocytes. (C) All sites shared. (D) Shared sites with a minimum absolute methylation difference of 5% or more between NEC and non-NEC and p<0.05 in both methods.

### Relationship Between DNA Methylation and RNA Transcription in NEC

We next explored the relationship between site-specific CpG methylation in our data and previously published RNA-Seq data obtained from a comparison of NEC colon and non-NEC control samples^19^. We evaluated the correlation between the directions of transcriptional and methylomic change between surgically resected NEC and non-NEC colon samples (Figure 6). Genes that contained differentially methylated CpG sites (p<0.05 and methylation difference > 10% in either direction) and that also encoded differentially expressed transcripts (p<0.05, LS Mean>=1) were identified. Within these, we identified 2,250 distinct genes representing 20,466 CpG sites (Table S2).

**Figure 6:**
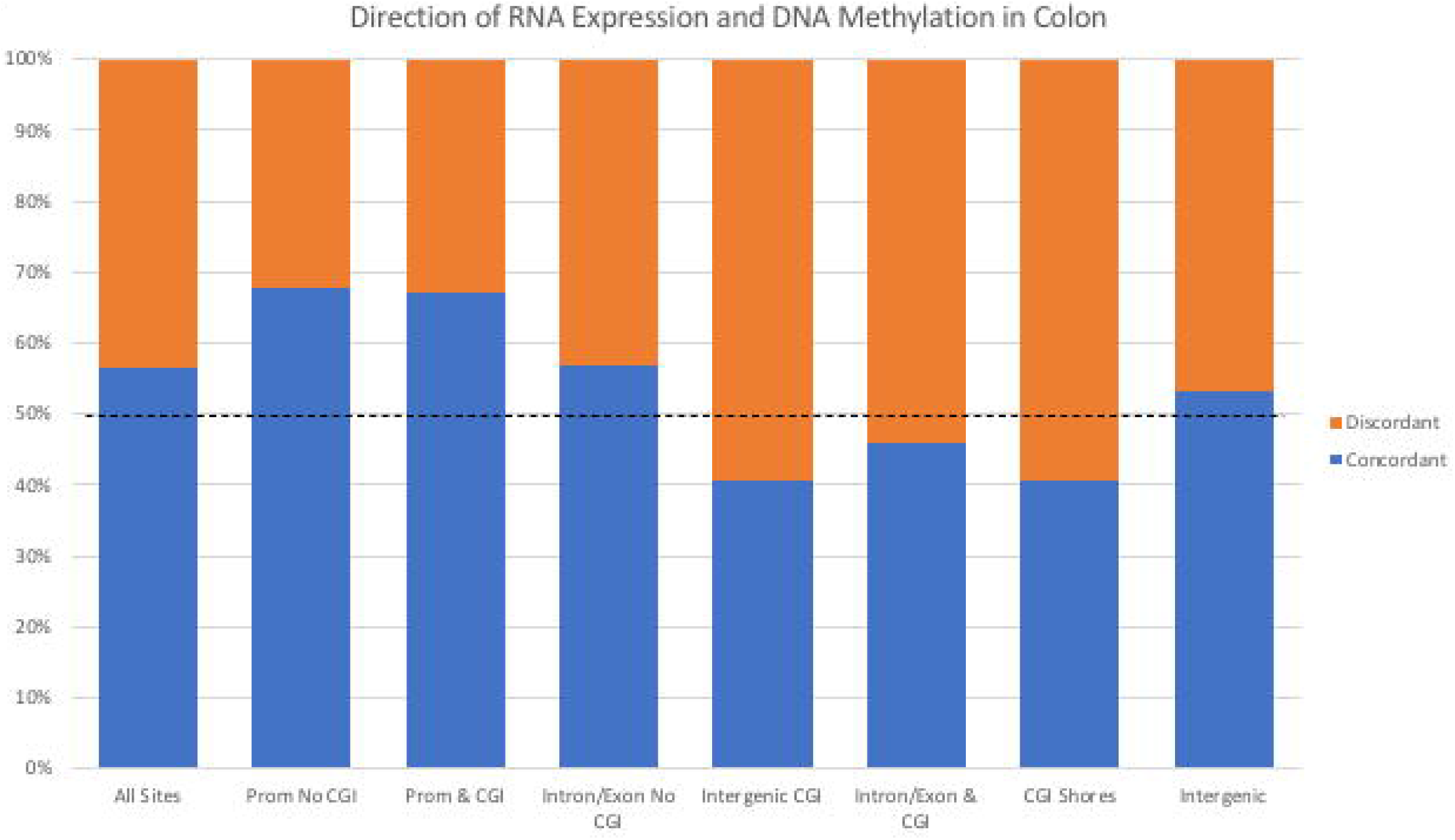
Relationship between RNA expression and DNA methylation by genomic element. Sites were considered concordant when NEC samples were hypermethylated and gene expression was lower in NEC samples and vice versa. Sites were considered discordant when NEC samples were hypomethylated and gene expression was higher in NEC samples and vice versa.

Among all CpG sites, 56.35% displayed DNA methylation changes that were inverse to the gene expression changes, which were designated as concordant. That is, their methylation increased when gene expression was <u>reduced</u> in NEC samples compared to non-NEC samples or vice versa. Similarly, 43.65% displayed directional DNA methylation changes that correlated with gene expression changes, which were designated as discordant. That is, their methylation increased when gene expression was increased in NEC samples compared to non-NEC samples or vice versa. We compared this distribution to sites within specific genomic elements using Pearson’s Chi-squared Test. We observed an increase in concordant sites in NEC compared to non-NEC samples in promoters outside CGIs (67.78%) and also within CGIs (67.23%). Similar trends were observed in introns/exons (56.81%) and intergenic regions not overlapping CGIs (53.20%). In contrast, we observed a reduction in concordant sites in NEC compared to non-NEC samples in CpG sites within CGIs. This was observed in intergenic CGI (40.66% concordant sites), intron/exon CGI (45.82%) and in CGI shores (40.64%). These trends reveal that genes whose expressions are modulated in NEC are more likely to display an inverse trend in DNA methylation change for CpG sites located outside of CGIs (or are within a promoter sequence). These genes (modulated in NEC) are more likely to display the same trend in DNA methylation change for CpG sites located inside CGIs or CGI shores (Table 4 and Figure 6).

**Table 4:** Relationship Between CpG Methylation and RNA Expression in NEC and non-NEC colon samples.

|  | Concordant | Discordant | Chi-square p-value |
| --- | --- | --- | --- |
| All Sites | 11533 (56.35%) | 8933 (43.65%) |  |
| Prom No CGI | 2524 (67.78%) | 1200 (32.22%) | p-value < 2.2e-16 |
| Prom & CGI | 316 (67.23%) | 154 (32.77%) | p-value = 3.175e-06 |
| Intron/Exon No CGI | 5861 (56.81%) | 4456 (43.19%) | p-value = 0.4523 |
| Intergenic CGI | 124 (40.66%) | 181 (59.34%) | p-value = 5.789e-08 |
| Intron/Exon & CGI | 947 (45.82%) | 1120 (54.18%) | p-value < 2.2e-16 |
| CGI Shores | 291 (40.64%) | 425 (59.36%) | p-value < 2.2e-16 |
| Intergenic | 1723 (53.20%) | 1516 (46.80%) | p-value = 0.0008285 |

### Gene-Specific Alterations in DNA Methylation Between NEC and Non-NEC Colon: Distribution and Pathway Analysis

The solution-phase hybridization approach we used for genome enrichment after bisulfite conversion allows the acquisition of high read-depth data within defined and relatively well characterized genomic regions. Therefore, CpG sites whose methylation is altered in a disease-specific fashion can be triangulated to their gene/region-specific location so that biological information regarding the underlying pathobiology of disease may be obtained.

Analysis of our data identified numerous differentially-methylated regions (DMRs) between NEC and non-NEC colon. Specifically, we identified 557 genomic loci containing gene bodies (introns/exons) or promoter regions with a difference in average methylation rate of at least 0.1 between NEC and non-NEC colon samples (Table S3). We further identified 4,367 differentially methylated single CpG sites (DMSs) between NEC and non-NEC colon samples using a p value, adjusted for multiple testing comparisons filter of q ≤0.10 (p value <1.72×10^−4^). Of these, we found 769 in exons, 2381 in introns, 822 in intergenic regions, 1154 in CGI shores and 12 in enhancers. Those located in CGIs numbered 527 and, of these, 85 of these were located within promoters, 172 in exons, 271 in introns, 108 in intergenic regions 1154 in CGI shores (Table S4).

Previous studies have demonstrated that DNA methylation signatures reveal considerable information regarding the molecular phenotype of the tissue/cell type(s) in question^10,20,21^. We therefore explored the functions of genes in which NEC-specific DMSs were identified using Ingenuity Pathways Analysis (IPA) software. We performed IPA analysis on genes whose promoters were found to contain altered CpG methylation in NEC colon versus non-NEC samples. We identified enrichment of genomic regions in numerous notable biological pathways under the control of a small number of known upstream regulators. For example, we found enrichment of genes in pathways including “FXR/RXR activation” (p=9.60×10^−9^), “LPS/IL-1 mediated inhibition of RXR function” (p=9.60×10^−9^) and “LXR/RXR activation (p=9.60×10^−9^) (Figure S2A-C and Table S5A). Predicted upstream regulators of these pathways include dexamethasone (p=1.37×10^−20^), HNF1A (p=4.63×10^−19^) lipopolysaccharide (p=8.02×10^−19^), TGFB1 (p=5.00×10^−17^) and TNF (p=7.70×10^−19^) (Table S5B).

Given that DNA methylation in CGI shores has been shown to be associated with transcription^19^, we also explored functional associations of genes containing differentially DMSs in CGI shores. We found that these were enriched in a number of pathways including “molecular mechanisms of cancer” (p=1.04×10^−16^), “human embryonic stem cell pluripotency” (p=8.86×10^−12^) and “axonal guidance signaling (p=7.39×10^−9^) (Figure S3A-C and Table S6).

We further performed IPA analysis of DMSs within CGIs whose DNA methylation levels were significantly altered between NEC and non-NEC colon (q≤0.05). We found that CpG sites within CGIs whose methylation levels were significantly altered in NEC versus non-NEC colon samples were located in genes that were enriched in pathways including “human embryonic stem cell pluripotency” (p=9.60×10^−9^), “regulation of the epithelial-mesenchymal transition pathway” (p=2.72×10^−8^) and “regulation of the epithelial-mesenchymal transition pathway in development” (p=6.62×10^−8^) (Figure S4A-C and Table S7A). Predicted upstream regulators of genes in these pathways included SOX2 (p=1.40×10^−13^), KMT2A (p=4.96×10^−11^), POU5F1 (p=1.03×10^−10^), KAT6A (p=4.38×10^−10^) and SHH (p=3.91×10^−9^) (Table S7B). Putative regulatory targets of these gene products are shown in Table S8.

### Putative NEC-Specific Regulatory Network Dysfunction

As described above, we found NEC-specific differential methylation in CGIs within defined canonical pathways. One putative upstream regulator identified as part of this analysis was POU5F1, which encodes a homeobox transcription factor and is located on chromosome 6. POU5F1 was predicted to regulate a number of downstream genes in our data (Table S8C). Significantly, we found that, according to previously published RNA-Seq data^19^, POU5F1 itself is differentially expressed in NEC colon compared to non-NEC control tissue. Given this, we further investigated whether predicted downstream POU5F1 target genes that were differentially methylated in the current study were also differentially expressed in surgical NEC. Using IPA, we identified 52 of these CGI-containing genes in our data. Of these, we found that 25 were also differentially expressed in NEC tissue compared to non-NEC controls. NEC-specific RNA expression and DNA methylation changes for these are shown in Table S8C. Given that a number of these genes themselves encode transcriptional regulators, we searched for evidence of functional interaction. As shown in Figure 7, we identified a number of putative regulatory associations between these genes. For example, there is evidence that WNT5A and FRZB, which are both integral to normal cell proliferation and development in the colonic crypt^22^ and are both altered in our NEC data at the epigenetic and transcriptional levels, are co-regulated by KLF4 which is also dysregulated in NEC. Reduced KLF4 expression was observed in NEC samples and WNT5A expression was also elevated in NEC, whereas FRZB expression was reduced. Notably, impaired Wnt pathway regulation leads to dysfunction of intestinal regeneration during necrotizing enterocolitis^23^. There is also evidence that KLF4 regulates the expression of TNFRSF25 whose expression is significantly upregulated in inflamed intestinal tissues^24^. Additionally, KLF4 may regulate the expression of the Snail Family Transcription Repressor 1 (SNAI1). Interestingly, SNAI2 gene expression is dramatically increased in NEC and a similar negative correlation with KLF4 expression has been observed in the epithelial-mesenchymal transition in colorectal cancer^25^.

**Figure 7:**
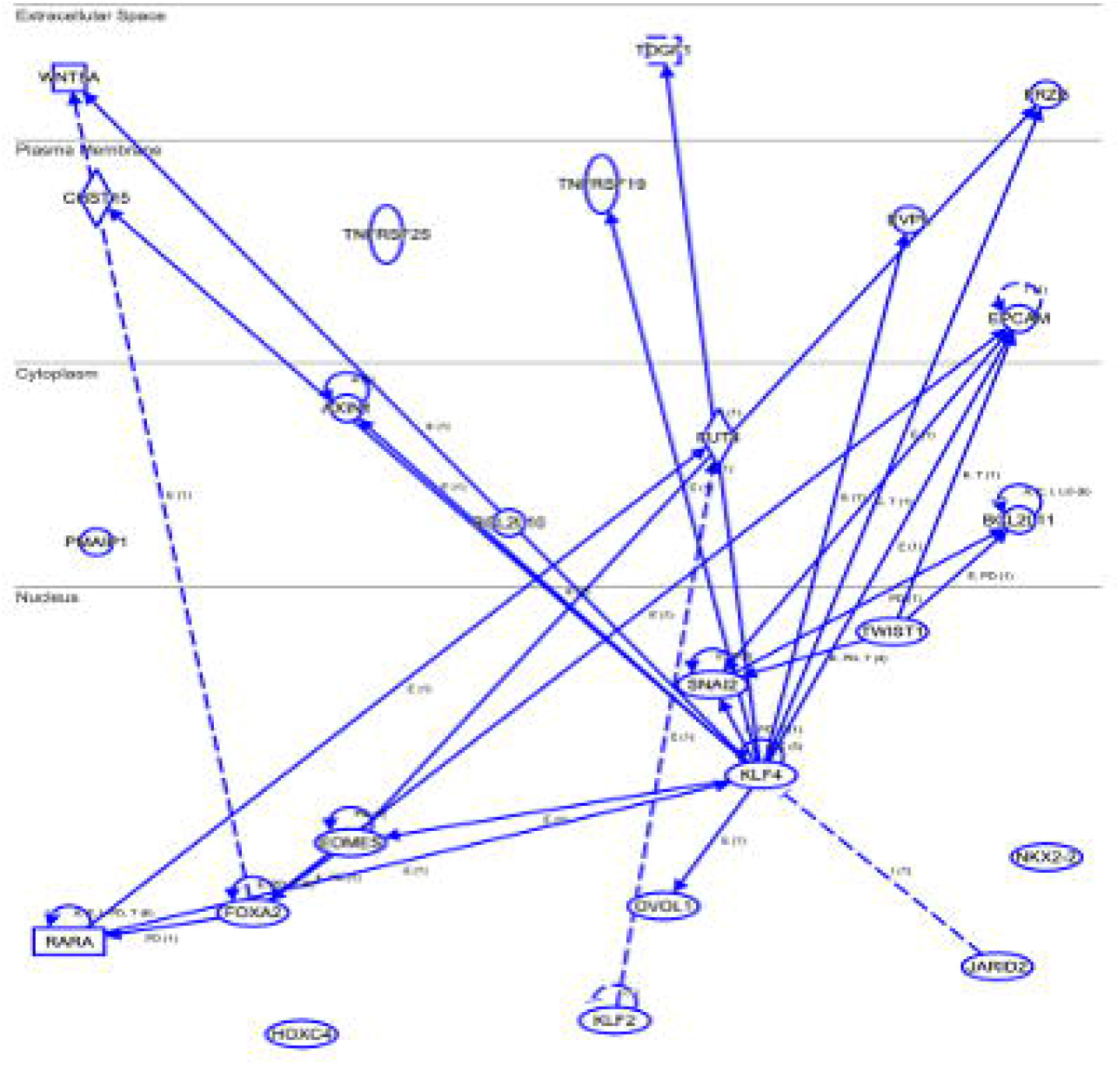
Putative regulatory associations for 25 POU5F1 target genes that were differentially methylated and differentially expressed in NEC versus non-NEC colon.

### NEC-Associated Hypermethylation is Evident in Stool Samples from Infants with NEC

To investigate the potential for DNA methylation signatures as biomarkers for the detection and phenotyping of NEC, we explored DNA methylation in stool samples from infants with NEC and normal controls (Table S9). As we previously observed in colonic samples, we found that CpGs in NEC stool samples were frequently more methylated than their premature control counterparts. As we observed in tissue, these effects were more pronounced in regions without CGIs, whereas in CGIs outside promoters, we found that DNA methylation was more balanced between NEC and control stool samples. CGIs within promoters displayed the least degree of hypermethylation in NEC, which is reminiscent of the tissue data in which these promoter CGIs were generally more methylated in non-NEC controls than NEC stool samples. Thus, as we found in tissue, the genomes of NEC stool samples exist in a hypermethylated state relative to their non-NEC counterparts, and this effect is most dramatically observed outside of CGIs (Figure 8 and Table 5).

**Table 5:** Proportion of CpG sites in different genomic elements that were hyper- or hypomethylated in NEC versus non-NEC control stool.

|  | Non-NEC | NEC | Chi-square p-value versus "All Sites" Distribution |
| --- | --- | --- | --- |
| All Sites | 493143 (28.97%) | 1209219 (71.03%) |  |
| Prom No CGI | 88555 (33.33%) | 177121 (66.67%) | p-value < 2.2e-16 |
| Prom & CGI | 123702 (42.91%) | 164558 (57.09%) | p-value < 2.2e-16 |
| Intron/Exon No CGI | 124088 (23.39%) | 406485 (76.61%) | p-value < 2.2e-16 |
| Intergenic CGI | 24037 (31.15%) | 53125 (68.85%) | p-value < 2.2e-16 |
| Intron/Exon & CGI | 119437 (33.28%) | 239401 (66.72%) | p-value < 2.2e-16 |
| CGI Shores | 21173 (32.91%) | 43170 (67.09%) | p-value < 2.2e-16 |
| Intergenic | 73345 (24.12%) | 230726 (75.88%) | p-value < 2.2e-16 |

**Figure 8:**
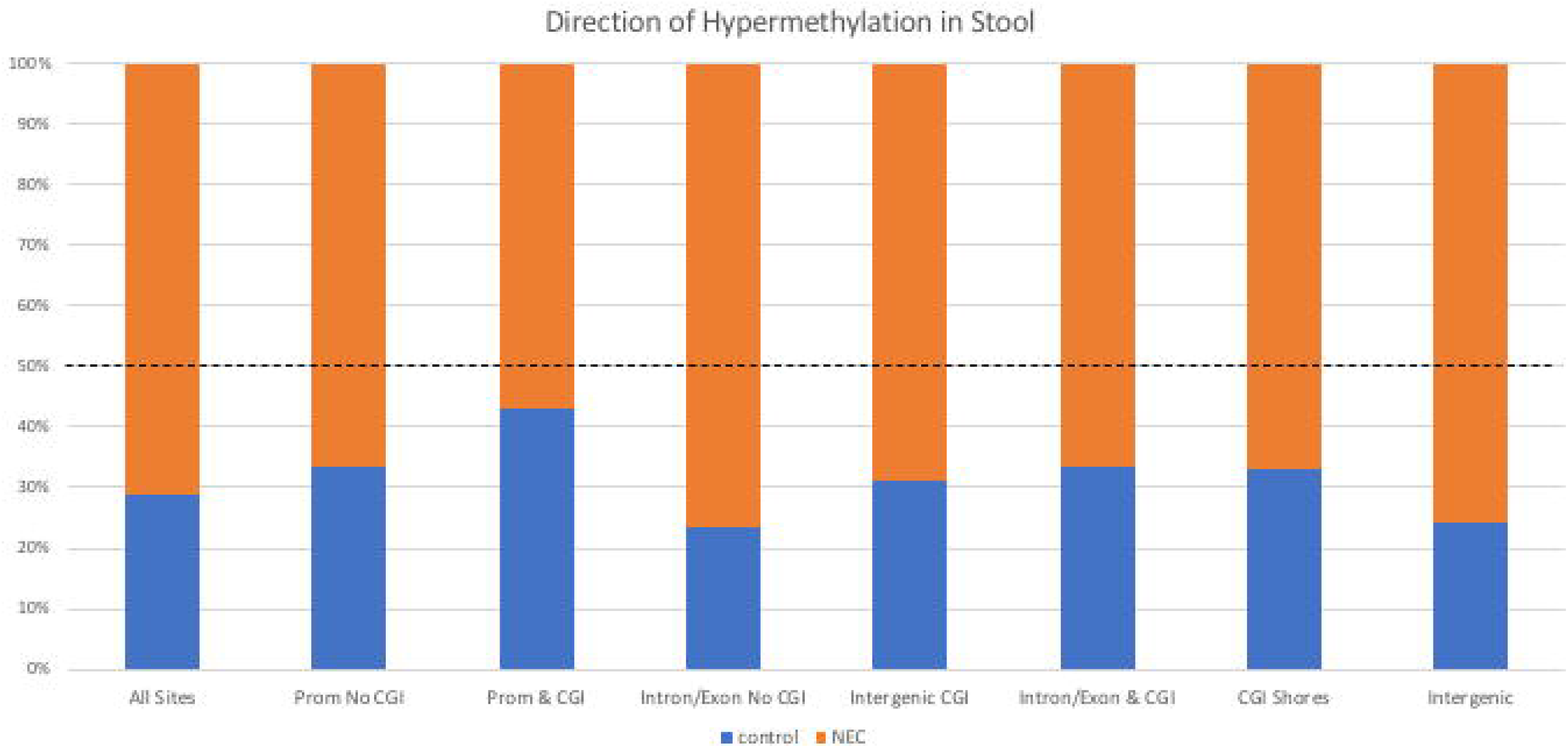
Proportion of CpG sites in different genomic elements that were hyper- or hypomethylated in NEC (orange) versus non-NEC control (blue) in stool. The dotted line represents a 50% ration of hyper- to hypo-methylated sites in a given genomic element.

Further examination of the association between the NEC tissue and stool data revealed a significant degree of correlation between NEC-associated DNA methylation signatures in both tissue and stool. Specifically, the correlation between CpG methylation levels of all targeted sites between NEC colon and stool was 0.980. For non-NEC controls, the value was 0.974 (Figure 9A). When we only considered sites with a methylation difference of >5% (in either way) and test p value <0.05 in both data set, the correlation coefficients were 0.849 between NEC samples and 0.724 between non-NEC samples (Figure 9B). When we compared the degree of CpG site-specific differential methylation between NEC and non-NEC for the two data sets, we found a correlation coefficient of 0.441 when all sites were considered (Figure 9C) and 0.524 when only sites with a methylation difference >5% between NEC and non-NEC (in either way) and test p value <0.05 were considered (Figure 9D). These results demonstrate that there is a positive degree of correlation between the impact of NEC on DNA methylation signatures in tissue and those in stool.

**Figure 9:**
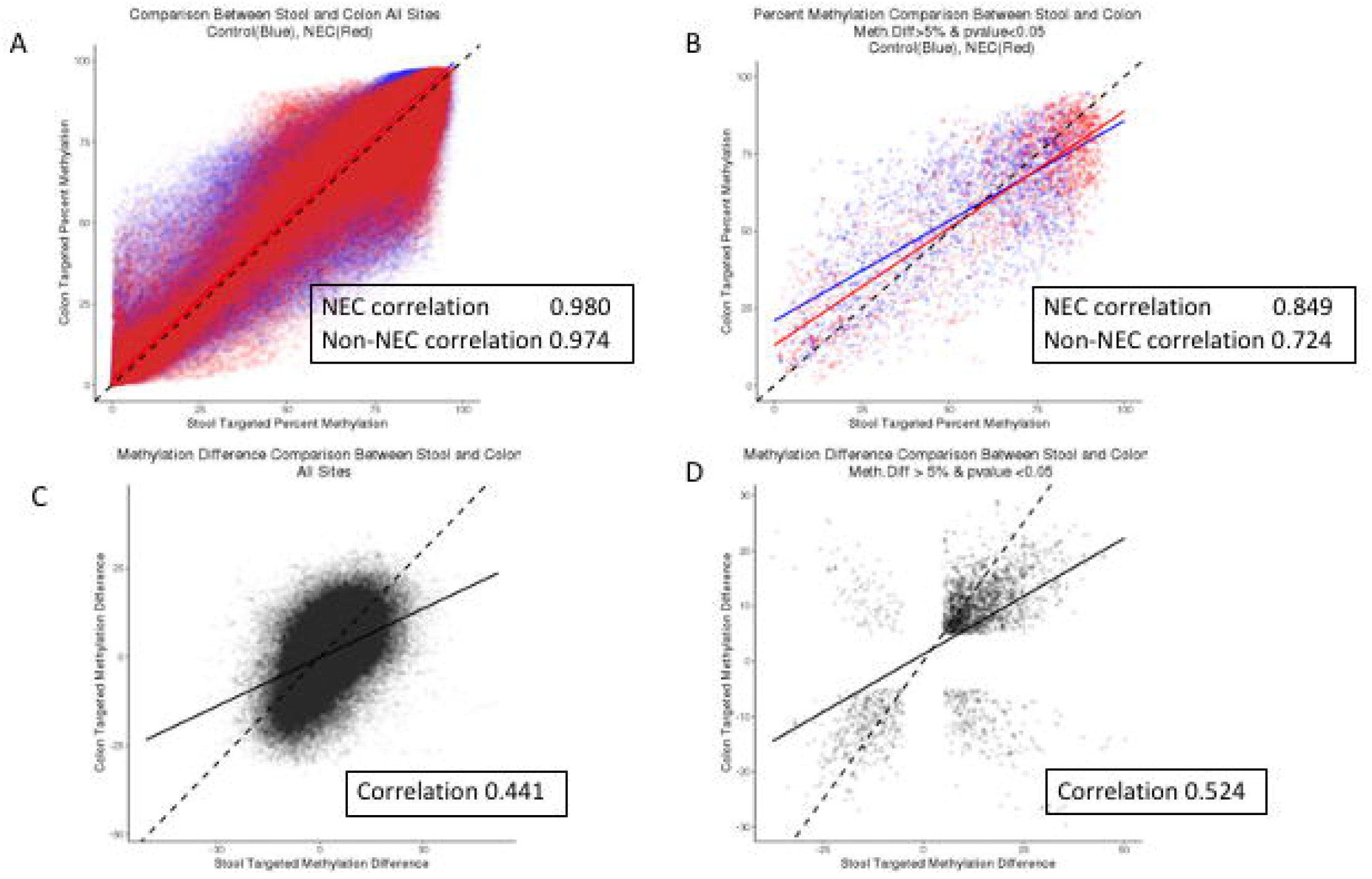
(A and B) Comparison of percent methylation between NEC (red) and non-NEC/control (blue) between colon tissue and stool. (A) All sites shared. (B) Shared sites with a minimum absolute methylation difference of 5% or more between NEC and non-NEC and p<0.05 in both methods. (C and D) Comparison of methylation difference (NEC versus non-NEC/control) between colon tissue and stool. (C) All sites shared. (D) Shared sites with a minimum absolute methylation difference of 5% or more between NEC and non-NEC and p<0.05 in both methods.

## Discussion

Healthcare providers in the neonatal intensive care unit are desperate for the discovery of a biomarker for NEC. By performing an initial broad overview in surgically resected intestine during active disease compared to a time when NEC had healed at reanastamosis, we discovered the methylome signature associated with the most severe form of the disease^19^. In these studies, we present a comprehensive evaluation of DNA methylation in NEC tissue samples compared to non-NEC controls. The most striking feature of our results is the significantly increased DNA methylation in samples from affected individuals. Of significance, these changes in methylation appear to be distributed unevenly between regions that either do or do not contain CGIs. CGI-containing regions display little or no significant skewing towards a hypermethylated state and this apparent resistance to NEC-associated change in methylation is most evident within those CGIs contained in promoter sequences. Therefore, it would appear that genomic regions associated with the transcriptional regulation of gene expression are less likely to display altered methylomes in the context of NEC. These observations suggest that the broad changes in methylation levels we observe are not the result of some indiscriminate process but, rather, are regulated by a disease-associated mechanism.

Notably, we found that a number of key elements of the DNA methylation machinery were themselves altered at the level of the epigenome in NEC. These epigenomic alterations were not associated with significant changes in the expression levels of the corresponding genes (data not shown) but this may be due to the fact that the tissue samples we assayed for methylation changes were obtained during surgical resection and therefore may well represent an epigenomic state that was influenced by earlier gene regulatory events, including altered expression of methylases (and related factors), that occurred closer to the onset of disease.

Some limitations of this case-control study are the lack of its prospective nature and the small sample size. Further investigation is required to determine the timing in which the hypermethylation occurs prior to the onset of NEC and importantly, what other clinical scenarios can lead to hypermethylation in the stool of infants. Furthermore, the method we employed for epigenomic analysis relied upon solution-phase hybridization of bisulfite converted DNA and next generation DNA sequencing. The DNA input requirements of this method precluded the use of single purified cell types and so, as a compromise, we extracted DNA from histological tissue sections that were largely confined to the colonic epithelium. One caveat of this approach is the likely presence of multiple cell types in any given sample, which of course will lead to variability in signal caused by differences in the relative dilution effect of these cell types. Therefore, we compared the current data to previously published whole-genome bisulfite sequencing data obtained using purified enterocytes from NEC and non-NEC control samples^19^. We were reassured to observe a high degree of correlation between these two datasets and we note that the previous findings in which we observed hypermethylation, particularly in colonic enterocytes from NEC samples, are evident in the current study. The current study significantly adds to the knowledge of NEC-associated epigenomic dysregulation because the targeted nature of the genomic analysis we employed enables high read-depth analysis within defined genomic regions as described herein.

Unsurprisingly, we found that DNA methylation changes within the colon during NEC are associated with directional changes in gene expression that vary according to genomic location. For example, we found that increased DNA methylation in promoter regions, regardless of whether or not the increase occurred within a CpG island, was more likely to be associated with reduced RNA expression of the corresponding genes. The opposite is also true, in that reductions in methylation within promotor regions (and CGIs in promoter regions) are more commonly associated with increased expression of the corresponding genes. DNA methylation changes in NEC compared to non-NEC samples that occurred in the same direction as expression of the corresponding gene were most frequently observed in intergenic CGI, intron/exon CGI and in CGI shores. These findings suggest that CGI methylation is associated with changes in gene expression and that these effects are influenced by whether or not the CGI resides in a promoter. Specifically, CGIs within gene promoters whose corresponding genes are upregulated in NEC are more likely to be less hypomethylated in NEC, whereas CGIs within intergenic regions and introns/exons are more likely to be hypermethylated.

Of particular interest, the trends in DNA methylation change that we identified in tissue samples also appear to be broadly evident in stool. Specifically, we found that CpG methylation in stool, as in tissue, is more frequently hypermethylated in NEC samples relative to non-NEC controls. In fact, although the genomic element-specific trends are approximately the same, the degree to which NEC samples are more frequently hypermethylated than controls was greater in stool than tissue. As in tissue, this phenomenon was less apparent within CpG islands, particularly those found in promotor regions. We also observed a significant degree of overall correlation between the tissue data and stool data. These results are important, because currently, there are no clinical blood or stool markers by which neonatologists and other healthcare practitioners can undertake surveillance of premature infants in the NICU to identify NEC in its early stages. This is problematic because the rapid onset of NEC can result in catastrophic inflammatory tissue damage. If the epigenomic changes we have identified in the stool and tissue samples of infants with NEC are also evident in early stages of disease, they may provide a desperately needed opportunity to develop noninvasive stool based screening tools for the early detection of NEC. The ability to diagnose NEC in its early stages would also enable the development of novel therapeutics that could be deployed before the overt manifestation of disease.

## Data Availability

Primary data are available from the authors on reasonable request.

## Figure Legends

**Figure S1:** Custom track (top) indicating methylation difference (NEC versus non-NEC) in colon. (A) DNMT3B gene. Region 1 shows NEC hypermethylation associated with CGI’s and promoter/enhancer region. Region 2: NEC hypomethylation in introns 4-6 overlapping CGI/shore region and enhancer element. (B) DNMT3B gene. NEC hypomethylation in promoter region. (C) TET1 gene. NEC hypomethylation in region 1 overlapping a promoter and CGI. NEC hypermethylation within exon 2 (region 2) and intron 3 (region 3). (D) TET3 gene. NEC hypomethylation in colon samples within a proximal promoter/enhancer region and also a distal enhancer. Regions of NEC hypermethylation were found within introns 1-3. (E) UHRF1 gene. NEC hypomethylation in the promoter region.

**Figure S2:** (A) Enrichment of genes in “FXR/RXR activation” (p=9.60×10^−9^) pathway. (B) Enrichment of genes in “LPS/IL-1 mediated inhibition of RXR function” (p=9.60×10^−9^) pathway. (C) Enrichment of genes in “LXR/RXR activation (p=9.60×10^−9^)” pathway.

**Figure S3:** (A) Enrichment of genes in “molecular mechanisms of cancer” (p=1.04×10^−16^) pathway. (B) Enrichment of genes in “human embryonic stem cell pluripotency” (p=8.86×10^−12^) pathway. (C) Enrichment of genes in “axonal guidance signaling” (p=7.39×10^−9^) pathway.

**Figure S4:** (A) Enrichment of genes in “human embryonic stem cell pluripotency” (p=9.60×10^−9^) pathway. (B) Enrichment of genes in “regulation of the epithelial-mesenchymal transition pathway” (p=2.72×10^−8^) pathway. (C) Enrichment of genes in “regulation of the epithelial-mesenchymal transition pathway in development” (p=6.62×10^−8^) pathway.

