## Supplemental Figures S1-S4 for "NEC-Associated DNA Methylation Signatures in Colon are Evident in Stool Samples of Affected Individuals"

Figure S1A

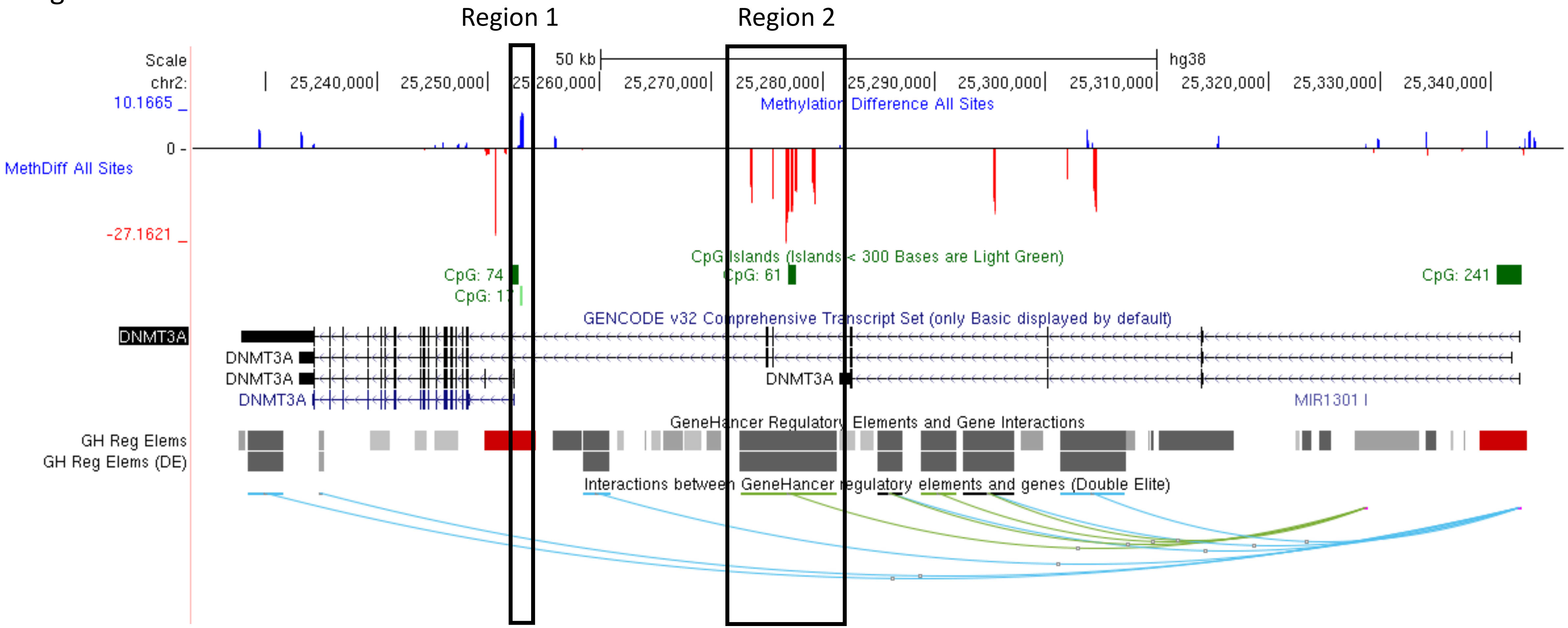
Region 1
Region 2
Scale
50 kb
hg38
chr2:
25,240,000
25,250,000
25,260,000
25,270,000
25,280,000
25,290,000
25,300,000
25,310,000
25,320,000
25,330,000
25,340,000
10.1665
Methylation Difference All Sites
0
MethDiff All Sites
-27.1621
CpG Islands (Islands < 300 Bases are Light Green)
CpG: 74
CpG: 17
CpG: 61
CpG: 241
GENCODE v32 Comprehensive Transcript Set (only Basic displayed by default)
DNMT3A
DNMT3A
DNMT3A
DNMT3A
DNMT3A
MIR1301
GeneHancer Regulatory Elements and Gene Interactions
GH Reg Elems
GH Reg Elems (DE)
Interactions between GeneHancer regulatory elements and genes (Double Elite)

Figure S1B

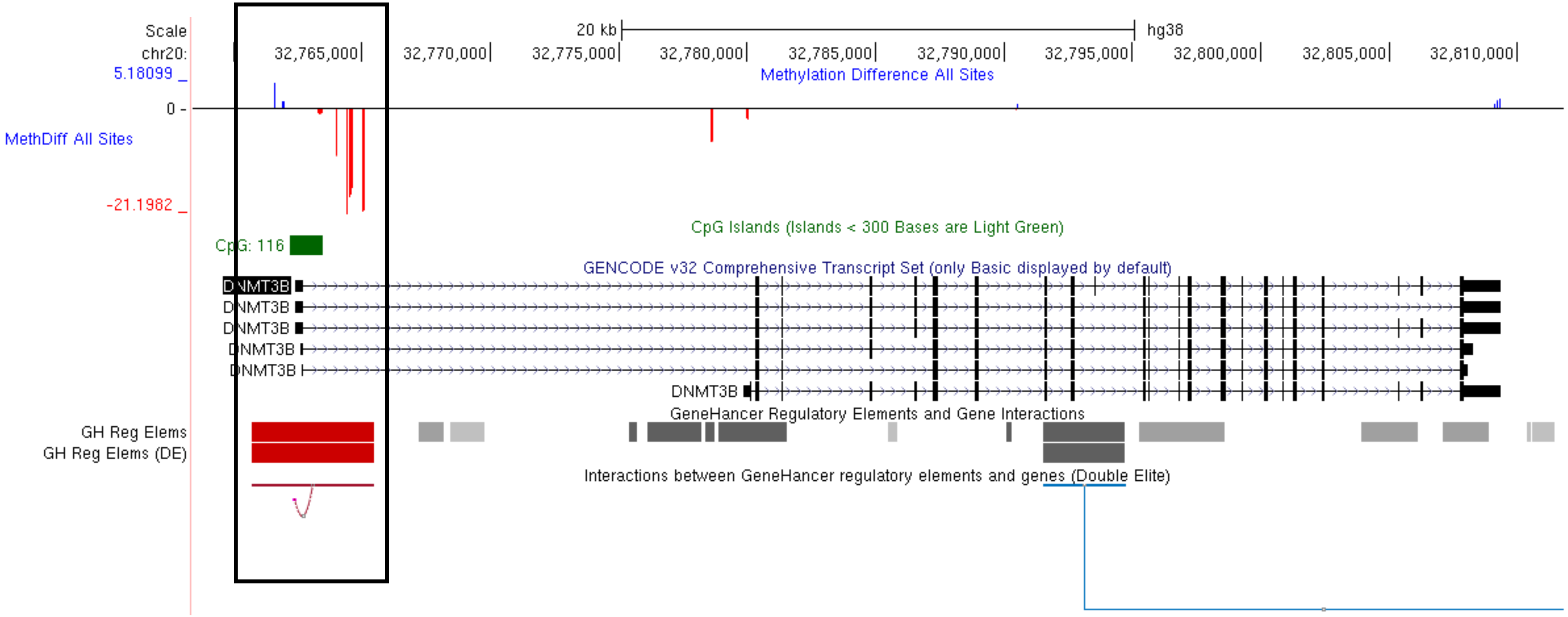
Scale
chr20:
20 kb
hg38
32,765,000
32,770,000
32,775,000
32,780,000
32,785,000
32,790,000
32,795,000
32,800,000
32,805,000
32,810,000
Methylation Difference All Sites
5.18099
0
-21.1982
MethDiff All Sites
CpG Islands (Islands < 300 Bases are Light Green)
CpG: 116
GENCODE v32 Comprehensive Transcript Set (only Basic displayed by default)
DNMT3B
DNMT3B
DNMT3B
DNMT3B
DNMT3B
DNMT3B
GeneHancer Regulatory Elements and Gene Interactions
GH Reg Elems
GH Reg Elems (DE)
Interactions between GeneHancer regulatory elements and genes (Double Elite)

Figure S1C

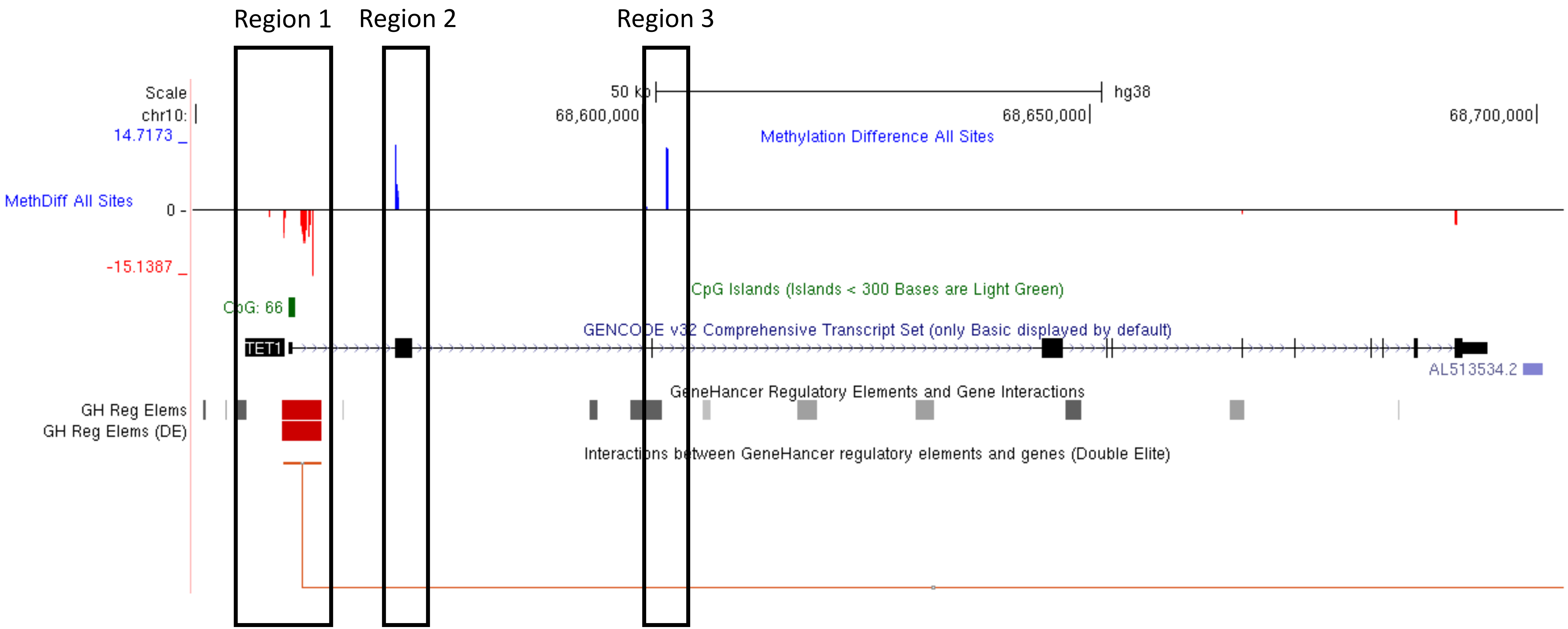
Region 1
Region 2
Region 3
Scale
chr10:
50 kb
hg38
68,600,000
68,650,000
68,700,000
14.7173 _
Methylation Difference All Sites
MethDiff All Sites
0 -
-15.1387 _
CpG Islands (Islands < 300 Bases are Light Green)
CpG: 66
GENCODE v32 Comprehensive Transcript Set (only Basic displayed by default)
TET1
AL513534.2
GeneHancer Regulatory Elements and Gene Interactions
GH Reg Elems
GH Reg Elems (DE)
Interactions between GeneHancer regulatory elements and genes (Double Elite)

Figure S1D

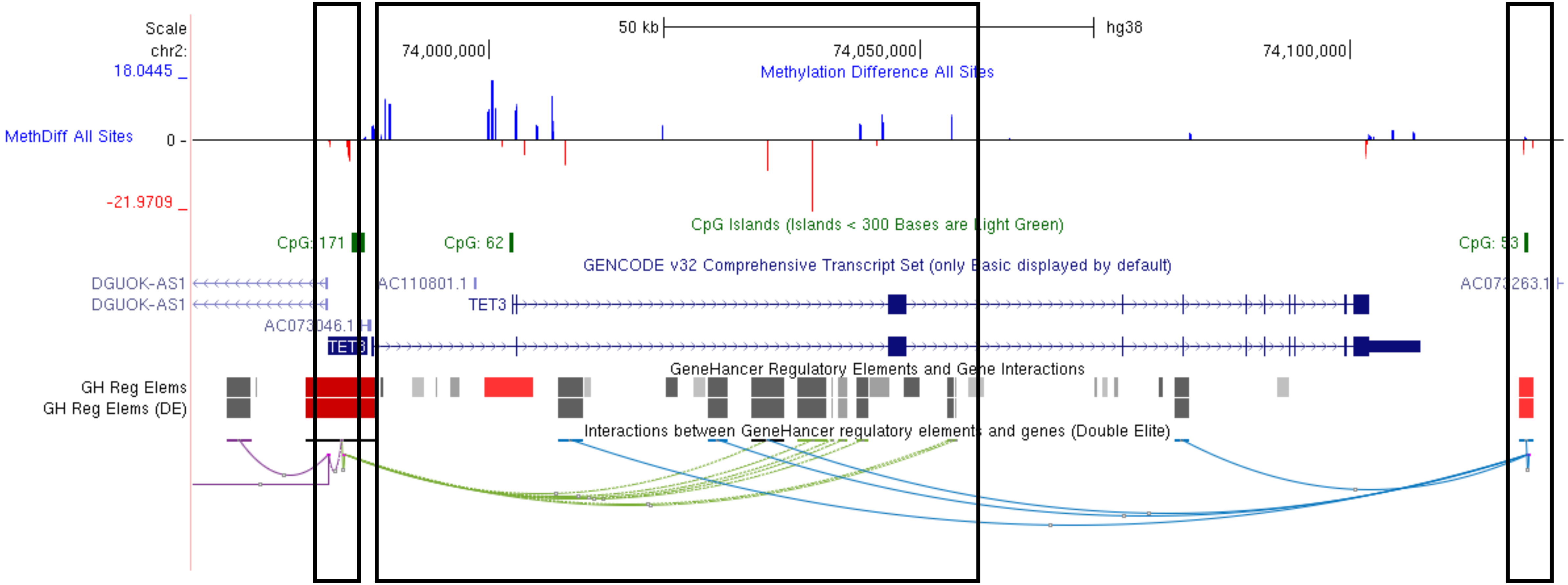

Scale
chr2:
50 kb
hg38
74,000,000
74,050,000
74,100,000
18.0445
Methylation Difference All Sites
MethDiff All Sites
0
-21.9709
CpG Islands (Islands < 300 Bases are Light Green)
CpG: 171
CpG: 62
GENCODE v32 Comprehensive Transcript Set (only Basic displayed by default)
DGUOK-AS1
DGUOK-AS1
AC110801.1
TET3
TET3
GeneHancer Regulatory Elements and Gene Interactions
GH Reg Elems
GH Reg Elems (DE)
Interactions between GeneHancer regulatory elements and genes (Double Elite)

Figure S1E

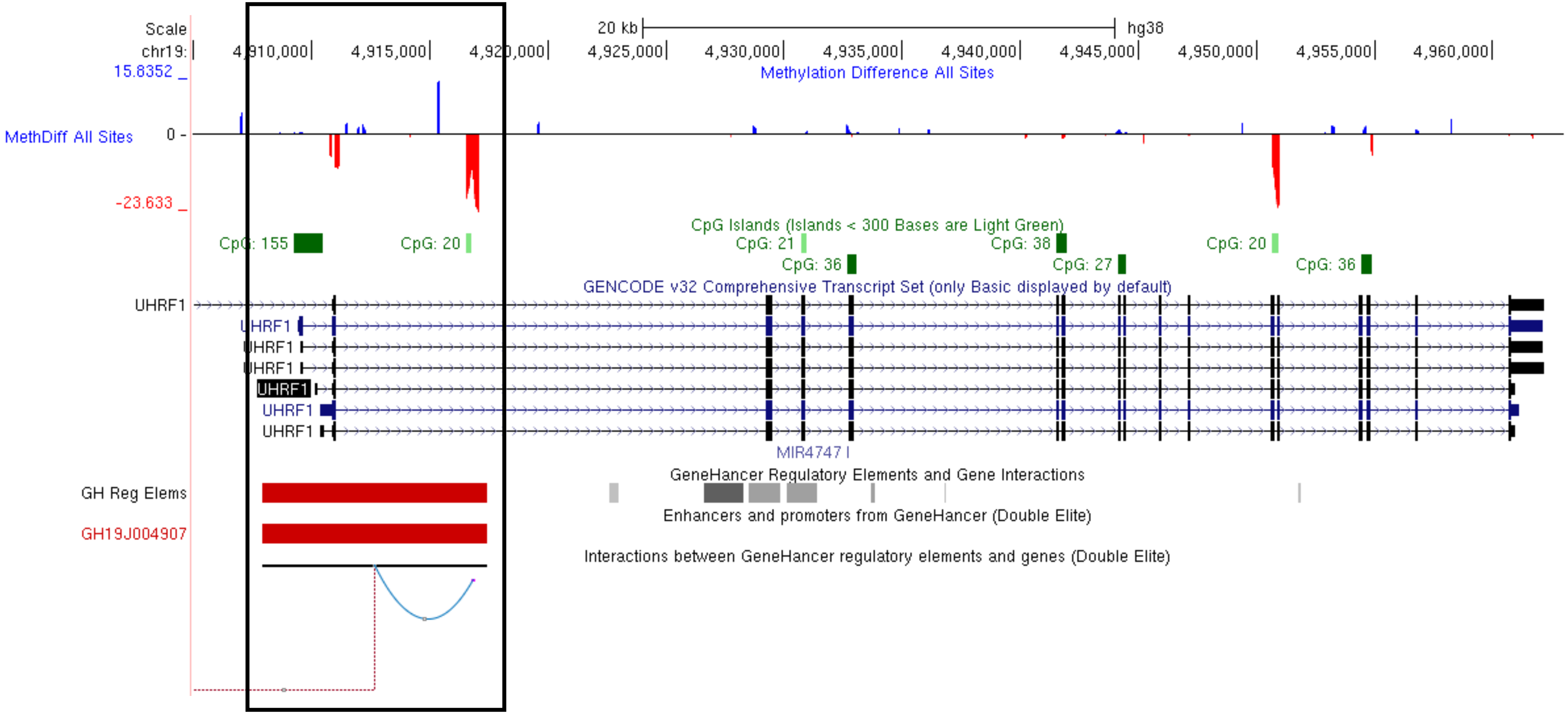

Scale
chr19:
20 kb
hg38
4,910,000
4,915,000
4,920,000
4,925,000
4,930,000
4,935,000
4,940,000
4,945,000
4,950,000
4,955,000
4,960,000
Methylation Difference All Sites
15.8352 _
MethDiff All Sites
0 -
-23.633 _
CpG Islands (Islands < 300 Bases are Light Green)
CpG: 155
CpG: 20
CpG: 21
CpG: 36
CpG: 38
CpG: 27
CpG: 20
CpG: 36
GENCODE v32 Comprehensive Transcript Set (only Basic displayed by default)
UHRF1
UHRF1
UHRF1
UHRF1
UHRF1
UHRF1
UHRF1
MIR4747
GeneHancer Regulatory Elements and Gene Interactions
GH Reg Elems
Enhancers and promoters from GeneHancer (Double Elite)
GH19J004907
Interactions between GeneHancer regulatory elements and genes (Double Elite)

### Figure S2A

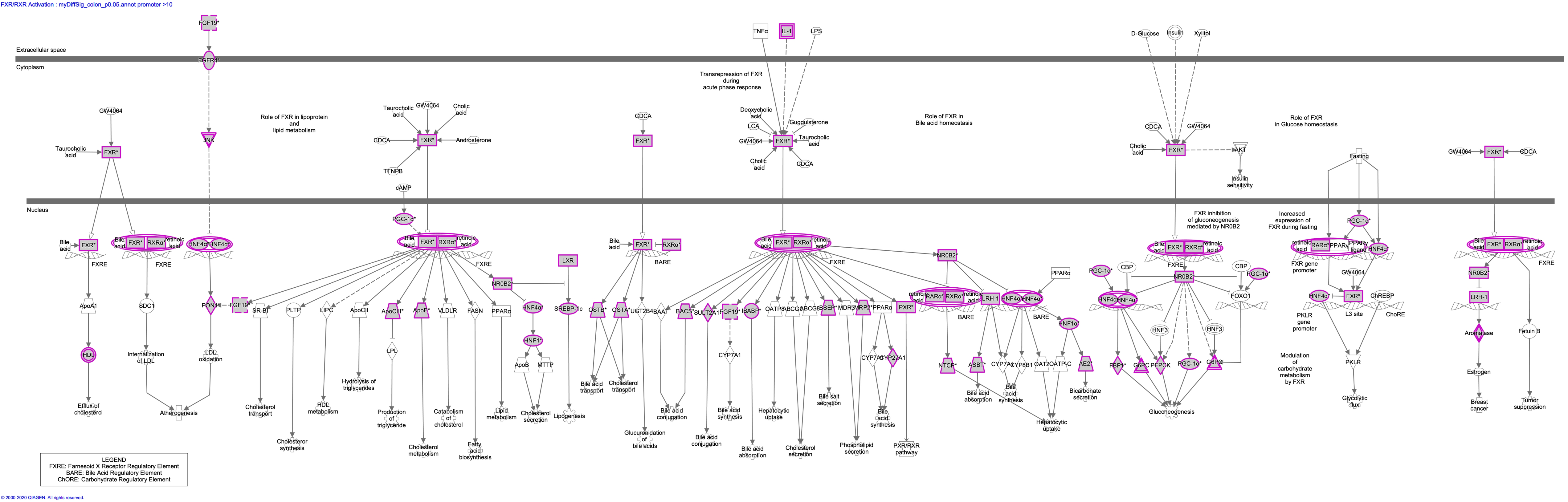


© 2000-2020 QIAGEN. All rights reserved.

LPS/IL-1 Mediated Inhibition of RXR Function : myDiffSig_colon_p0.05.annot promoter >10

### Figure S2B

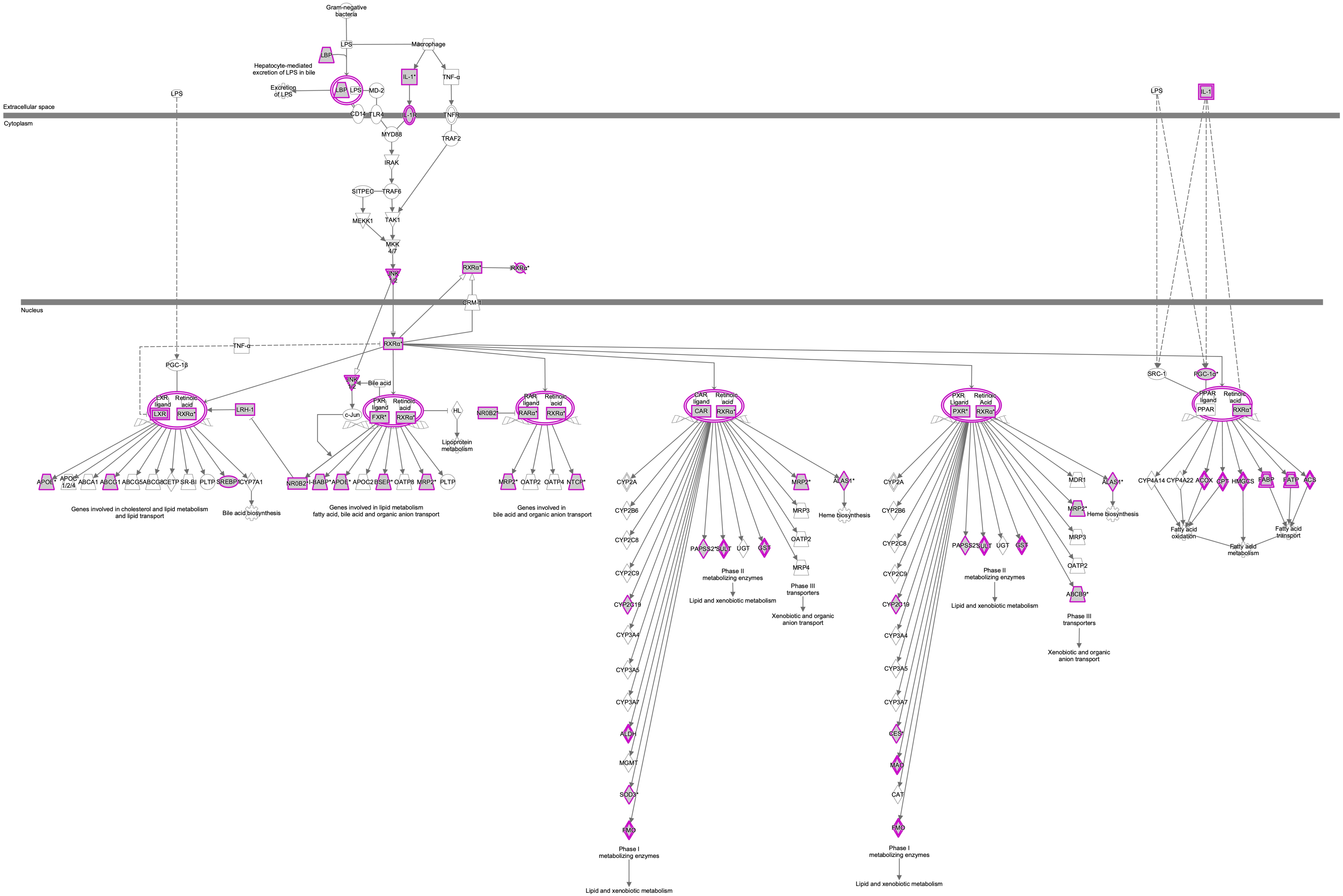


© 2000-2020 QIAGEN. All rights reserved.

LXR/RXR Activation : myDiffSig_colon_p0.05.annot promoter >10

### Figure S2C

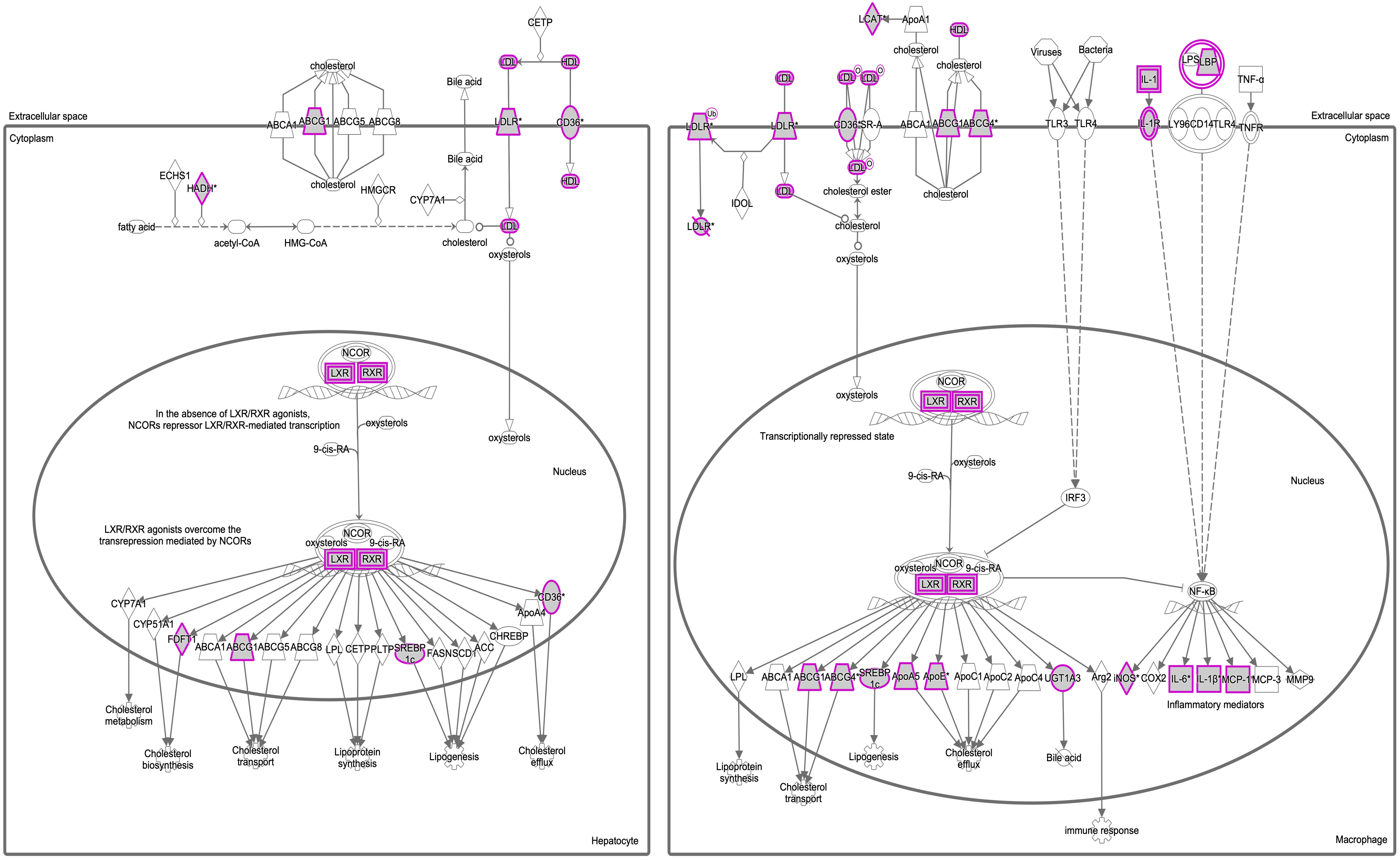


© 2000-2020 QIAGEN. All rights reserved.

Molecular Mechanisms of Cancer : myDiffSig_colon_p0.05.annot shores >5

### Figure S3A

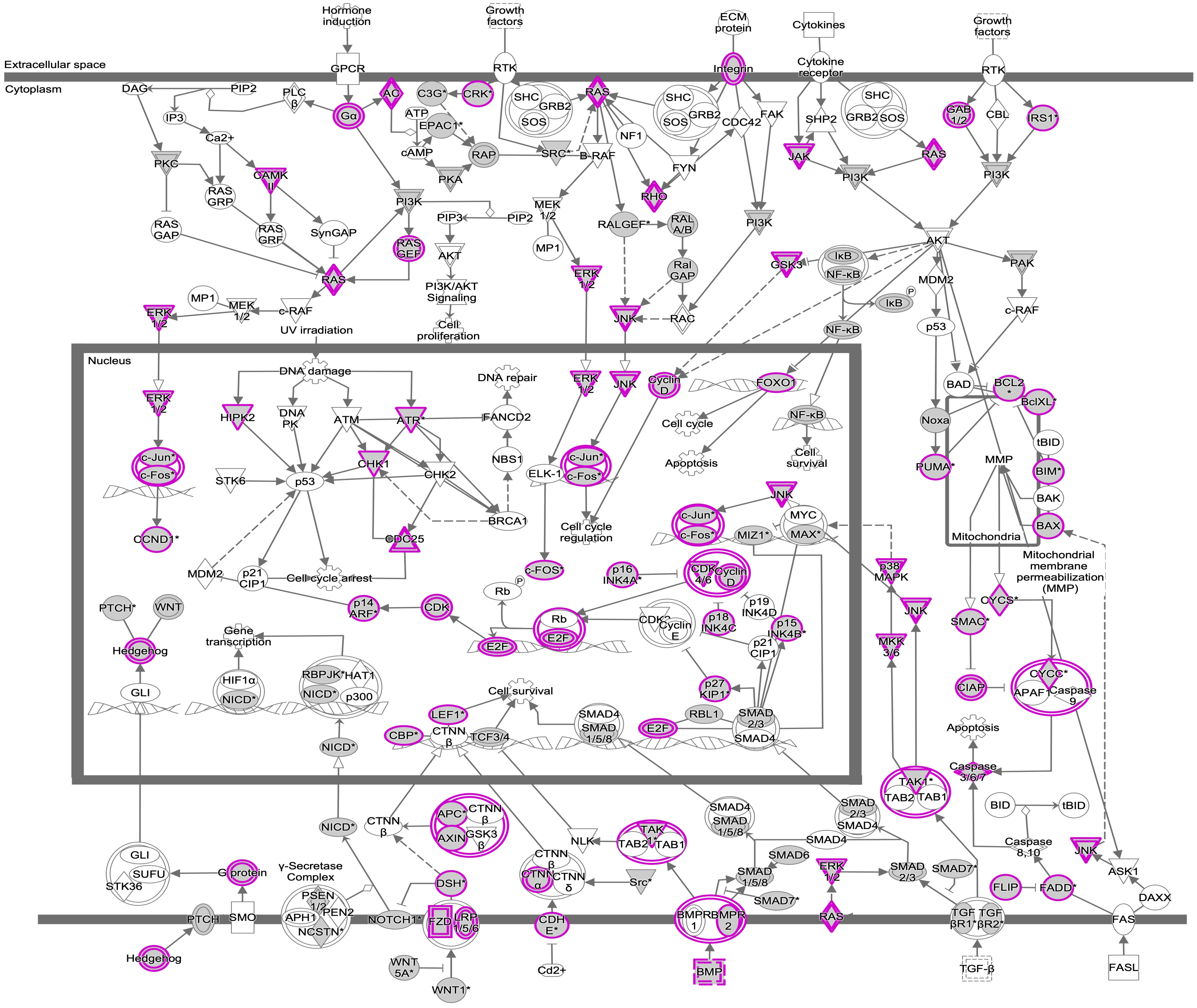


© 2000-2020 QIAGEN. All rights reserved.

Human Embryonic Stem Cell Pluripotency : myDiffSig_colon_p0.05.annot shores >5

Figure S3B

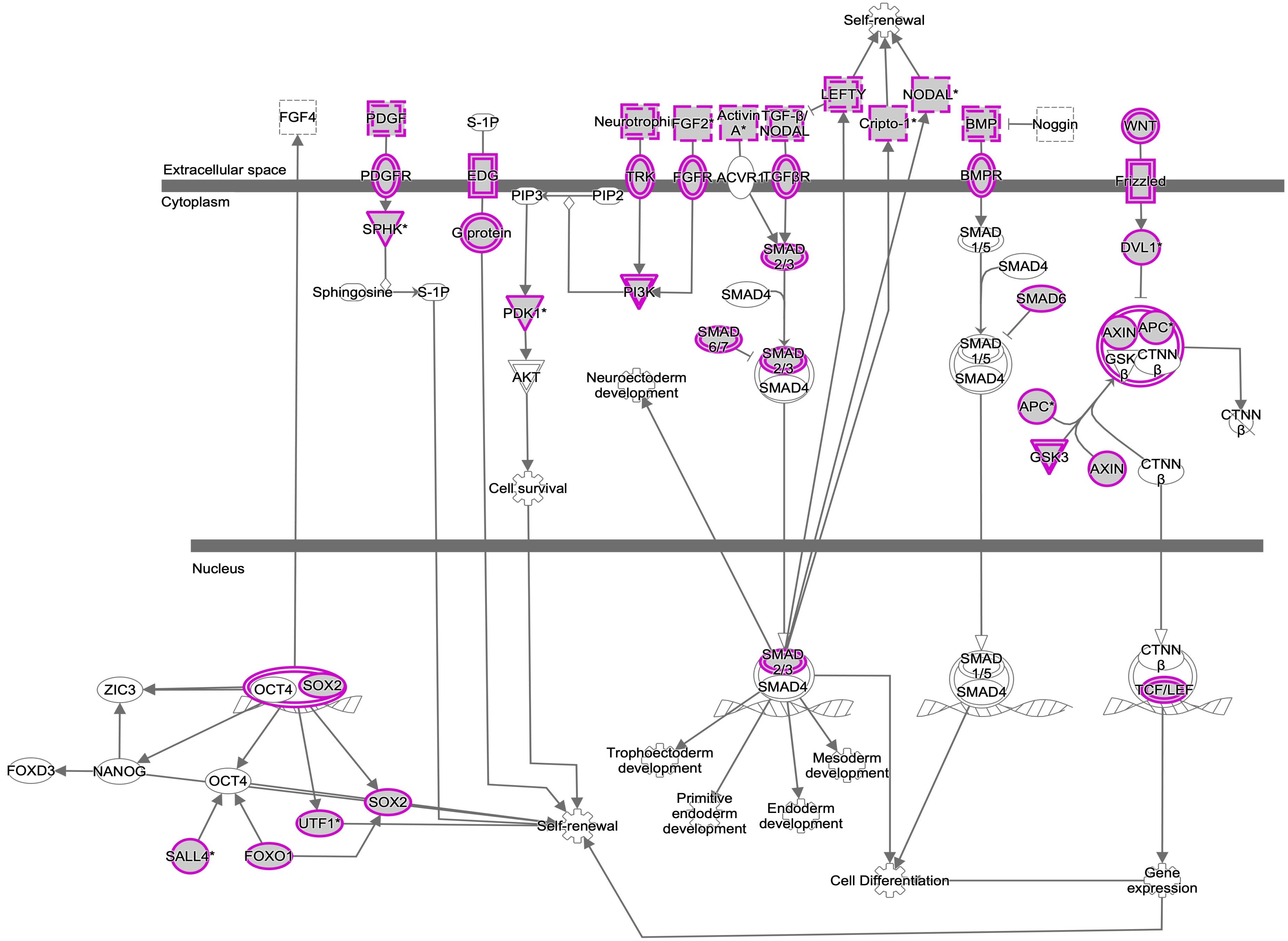


© 2000-2020 QIAGEN. All rights reserved.

### Figure S3C

Axonal Guidance Signaling : myDiffSig_colon_p0.05.annot shores >5

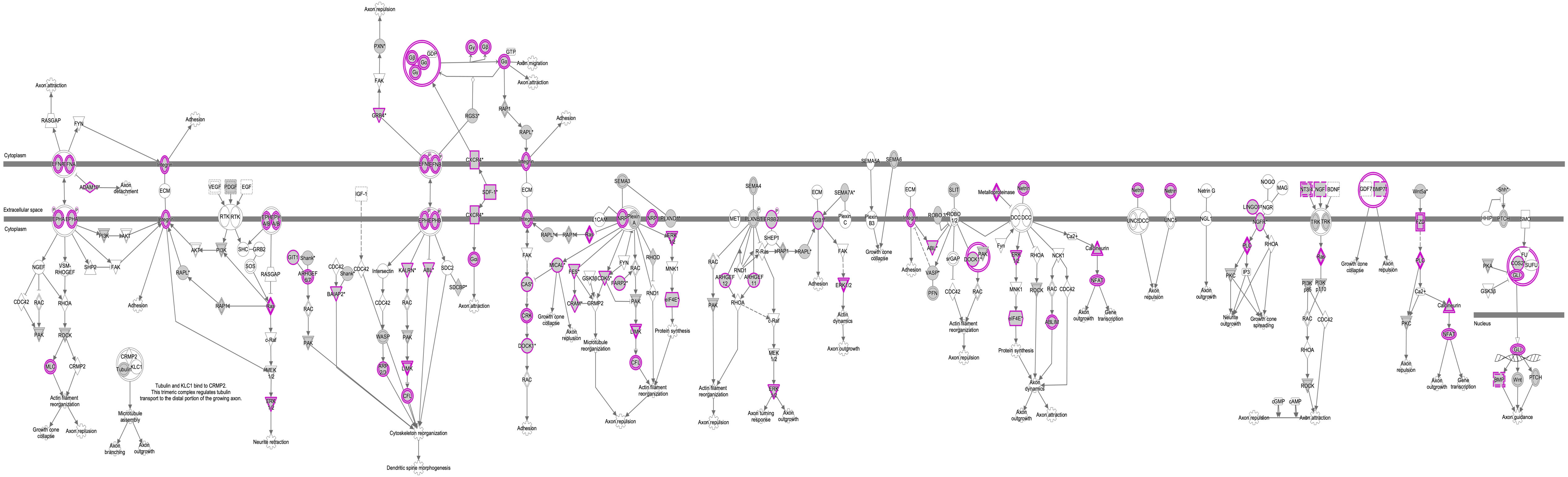

© 2000-2020 QIAGEN. All rights reserved.

### Figure S4A

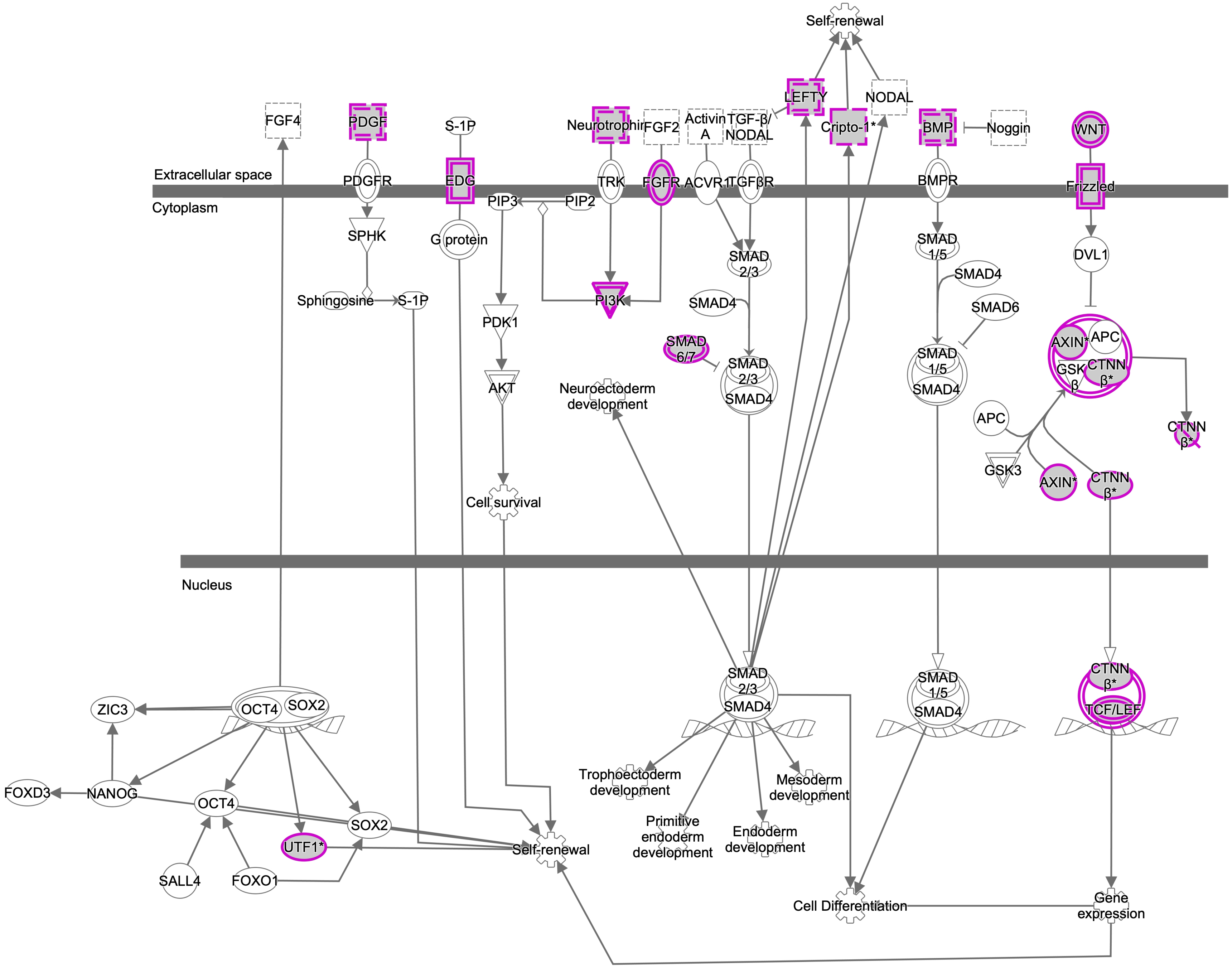


© 2000-2020 QIAGEN. All rights reserved.

### Figure S4B

Regulation of the Epithelial-Mesenchymal Transition Pathway : myDiffSig_colon_p0.05.annot CGI >5

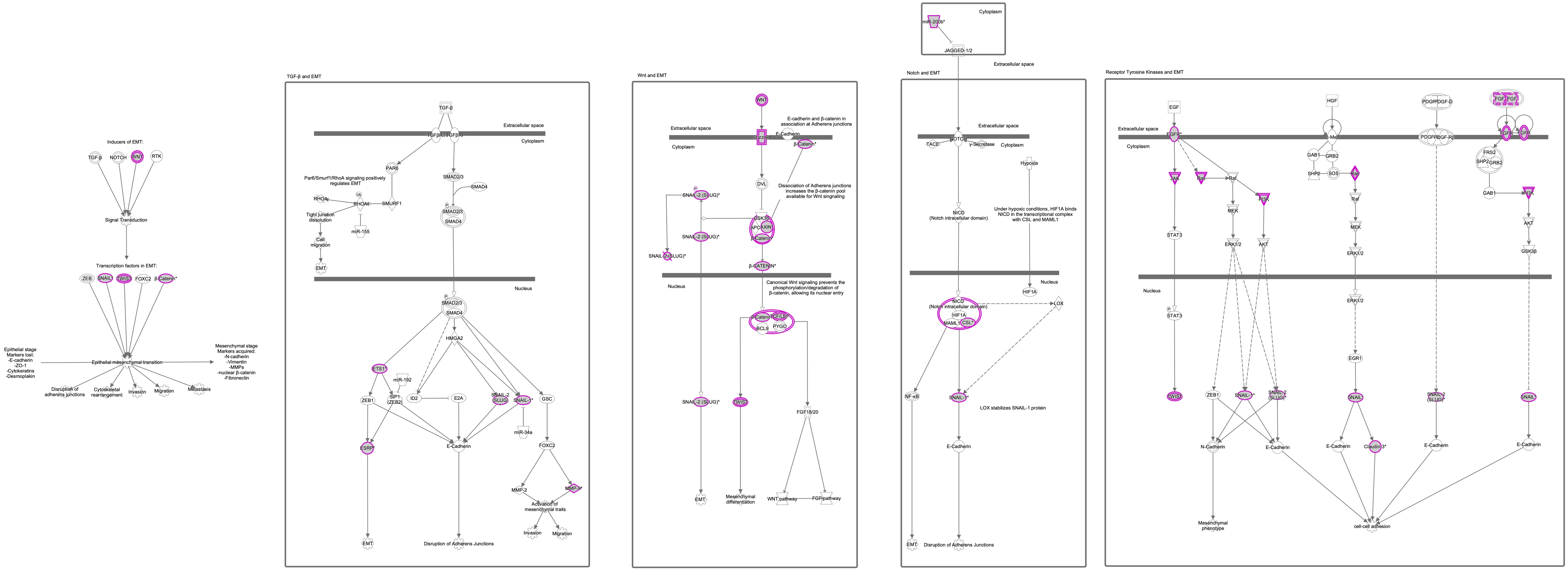


© 2000-2020 QIAGEN. All rights reserved.

### Figure S4C

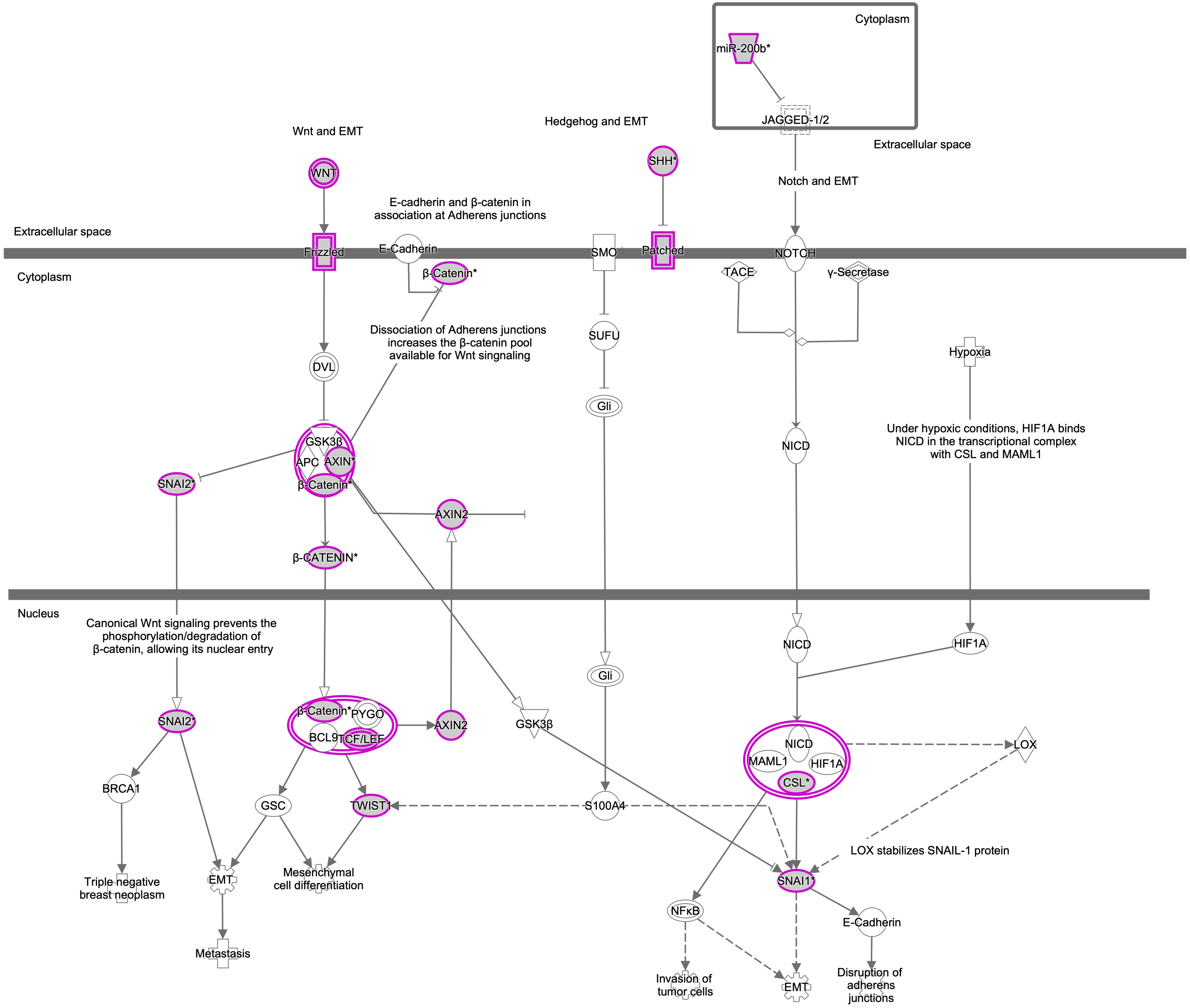


© 2000-2020 QIAGEN. All rights reserved.
